# Pathway analysis identifies novel non-synonymous variants contributing to extreme vascular outcomes in Williams-Beuren syndrome

**DOI:** 10.1101/2022.09.21.22280107

**Authors:** D. Liu, C.J. Billington, N. Raja, Z.C. Wong, M.D. Levin, W. Resch, C. Alba, D.N. Hupalo, E. Biamino, M.F. Bedeschi, M.C. Digilio, G.M. Squeo, R. Villa, P.C.R. Parrish, R.H. Knutsen, S. Osgood, J.A. Freeman, C.L. Dalgard, G. Merla, B.R. Pober, C.B. Mervis, A.E. Roberts, C.A. Morris, L.R. Osborne, B.A. Kozel

## Abstract

Supravalvar aortic stenosis (SVAS) is a characteristic feature of Williams-Beuren syndrome (WBS). SVAS is present in 67% of those with WBS, but severity varies; 21% have clinically significant SVAS requiring surgical intervention while 33% have no appreciable aortic disease. Little is known about genetic modifiers outside the 7q11.23 region that might contribute to SVAS severity. To investigate, we collaboratively phenotyped 473 individuals with WBS and performed the largest whole-genome-sequencing study to date. We developed a set of strategies for modifier discovery including extreme phenotyping (surgical SVAS vs. no SVAS) and prioritization of non-synonymous variants with increased predicted functional impact along with an allele frequency difference between the extreme phenotype groups. We identified pathways enriched in common or less frequent variants, followed by association testing of SVAS severity with the enriched pathways. The common variant analysis identified pathways including the extracellular matrix and the innate immune system, while pathways encompassing adaptive immunity, ciliary function, lipid metabolism and PI3KAKT were captured by both the common and less frequent variant analyses. Cell cycle and estrogen responsive pathways were among those identified through the less frequent variant analysis. Among the 69 genes reported in other large genome wide association studies assessing aortic traits, 11 genes, including *PCSK9* and *ILR6,* were found in our study, suggesting overlapping disease mechanisms. In summary, this study presents novel strategies for identification of disease modifiers in rare conditions like WBS.

**Graphical Abstract:** 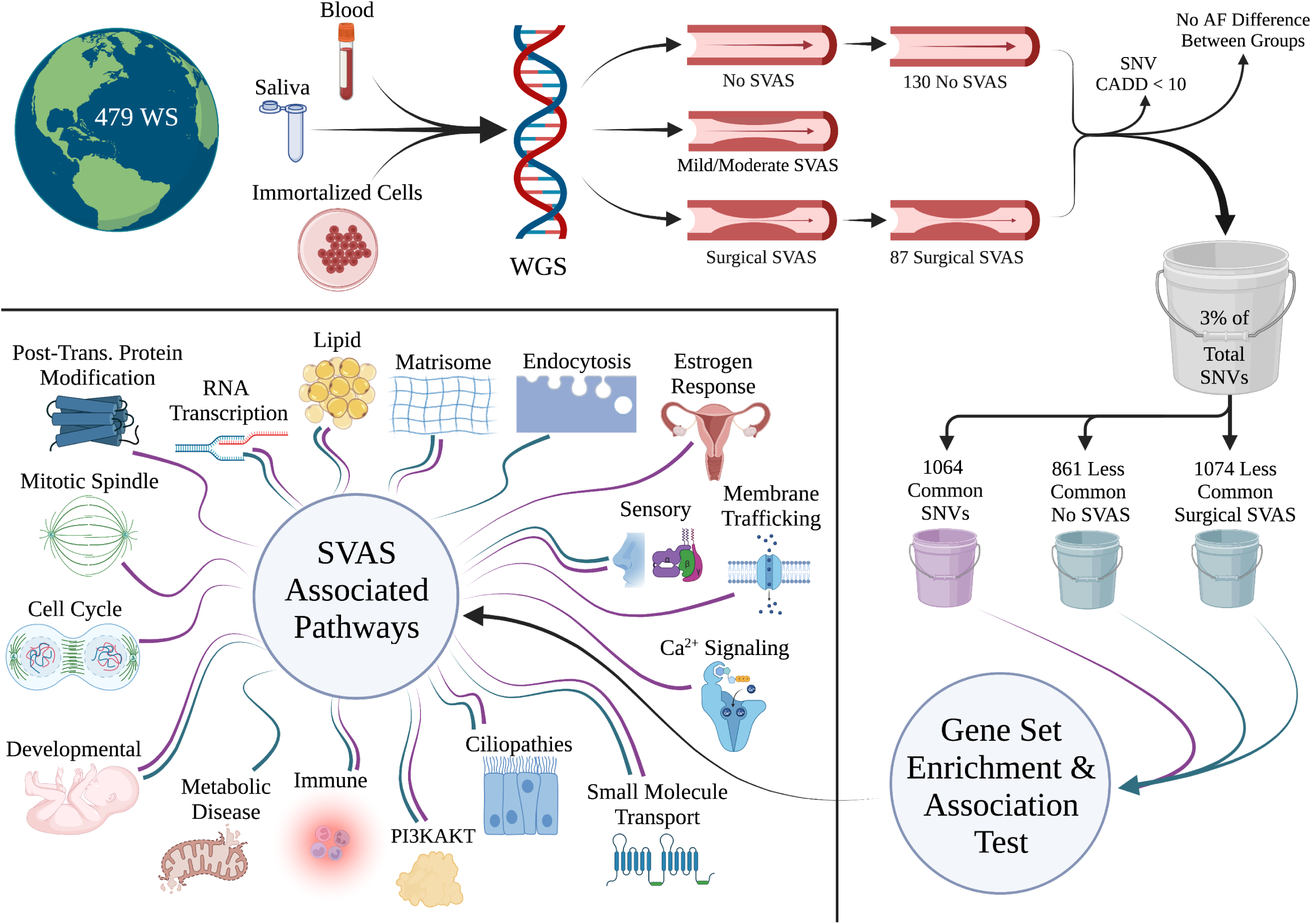

## Introduction

Williams-Beuren syndrome (WBS), caused by deletion of 1.5-1.8 Mb on human 7q11.23, is a multi- system disorder characterized by distinctive facies, a typical neurodevelopmental profile and cardiovascular disease^1^. It occurs in 1 of 7,500 individuals^2^ and is *de novo* in almost all cases. The cardiovascular disease in WBS is mediated by the deletion of elastin (*ELN*) from this region^3–7^ and consists of focal large and medium artery stenosis in the setting of a more global decrease in arterial caliber. Supravalvar aortic stenosis (SVAS), which is the focal narrowing of the ascending aorta above the aortic valve, is the feature most commonly associated with WBS^8–10^. Although more than 95% of individuals with WBS share the same basic deletion on chromosome 7q11.23, their outcomes for SVAS vary: about 20% have clinically significant SVAS requiring surgical intervention in infancy or childhood; in contrast, 30-40% of individuals with WBS never develop SVAS^9, 11, 12^. It has been unclear what features (genetic or otherwise) predispose to these extreme outcomes.

Genome-wide association (GWAS) studies have allowed investigators to connect genetic variation with an increasing number of traits and diseases. The impact of these types of studies has recently been amplified by the availability of genomic datasets from the UK BioBank^13, 14^, NIH All of US^15^, and TOPMED^16^. These databases hold genomic information on hundreds of thousands of individuals and have enabled discoveries on a myriad of common health conditions^17, 18^ and quantitative traits.

Although individually uncommon, together rare diseases impact 10% of the population in the United States (source: https://ncats.nih.gov/news/events/rdd). However, few of the nearly 10,000 GWAS reported in the NHGRI-EBI GWAS Catalog (source: https://www.ebi.ac.uk/gwas), were performed on cohorts of individuals with rare conditions. The application of GWAS to rare conditions such as WBS has been challenging because most existing GWAS methods and statistical packages were developed for studies with thousands of participants, which is unattainable for most rare disease studies, including WBS. Likewise, techniques to improve power, such as paired eQTL analysis^19^ of affected tissues, are challenging in difficult-to-access tissues like the aorta. As such, existing studies in rare *de novo* conditions, like WBS, have primarily focused on correlation of phenotype with variants within the disease-specific locus or region^20, 21^. Therefore, alternative analytical strategies are needed for the study of rare diseases using whole genome sequencing (WGS) data.

To search for genetic modifiers outside the 7q11.23 region contributing to SVAS severity with limited sample size, we proposed a set of strategies centered on the question of whether individuals with extreme SVAS phenotypes (surgical SVAS and no SVAS) exhibit a burden of non-synonymous variants (hereafter variants) that are enriched in a small number of biological pathways. The concept of pathway enrichment, which has been widely used in mRNA expression studies^22^, has been recently incorporated into GWAS analysis^11, 23, 24^. Including pathway information in GWAS analysis may be critical to identifying missing heritability^25, 26^. Due to the small sample size in our WBS study, we aimed to control false discovery of pathways associated with extreme phenotypes. One crucial prior step is to pre-select the most influential variants--those with greater likelihood of a functional impact as measured by combined annotation dependent depletion (CADD) score^27^, and variants with allele frequency (AF) differences between the surgical SVAS and no SVAS groups (either more than 5% difference in AF for common variants or variants present with AF > 1% in one extreme group and absent in the other--for a total of three separate analyses). This filtration step was intended to dramatically reduce the total number of variants to be considered for downstream pathway enrichment by genes with common variants or less frequent variants^17, 28^. We then tested association of SVAS severity with enriched pathways^29^ for identifying association of rare^30, 31^ and common variants^23, 32^. The last part of our strategy included consolidating the pathways with a large portion of overlapping genes to one pathway and comparing the genes on the consolidated pathways to those reported by large GWAS studies on aortic traits and diseases in the NHGRI-EBI GWAS catalog.

In this study, we applied these proposed strategies to the whole-genome sequencing (WGS) data generated from 225 participants with extreme phenotypes from the overall 473-member WBS cohort, contributed from four international groups for the largest WGS investigation in WBS to date. The work here expands on our previous exome-based pilot study^11^, with paired goals of replicating common-variant findings in the larger group and growing our analysis to evaluate pathways enriched by less frequent variants. By defining the pathways driving differences in SVAS outcome, we offer insight into the mechanism of elastin insufficiency mediated disease and hope to highlight potential new pathways that can be targeted by rational therapeutics. Additionally, we provide proof of principle that the combination of strategies used here can be applied to a range of rare diseases.

## Methods

### Consent

All participants alive at the time of enrollment or their caregivers signed informed consent forms to participate in research that included genome sequencing. One hundred and eighty participants signed consent approved by the NIH Institutional Review Board (IRB) through the WBS DNA and Tissue Bank (NCT02706639), 197 signed consent approved by the Reno Institutional Review Board of the University of Nevada, 20 signed the consents approved by the University of Toronto Health Sciences Research Ethics Board (those 217 were shared under the umbrella of the Nevada-Toronto collaboration- the Nevada samples had been previously de-identified and transferred to U Toronto), 10 signed consent approved by the Boston Children Hospital Internal Review Board, and 64 consented to participate in the Telethon Biobank in Italy and approved by Fondazione IRCCS Casa Sollievo della Sofferenza Ethics Board. Two additional NIH samples were derived from tissue donated after death and were considered exempt. The data were analyzed under the NIH approved protocol.

The NIH IRB, the University of Toronto Health Sciences Research Ethics Board, the Boston Children Hospital Ethics Board, Fondazione IRCCS Casa Sollievo della Sofferenza Ethics Board and the University of Nevada Reno Institutional Review Board gave ethical approval for DNA collection and genetic investigation. The NIH IRB gave ethical approval for the combined analysis of the de-identified samples.

### Procedures used in whole genome sequencing data processing

Samples were collected in one of three forms: whole blood, saliva, or immortalized cell lines derived from blood. DNA was prepared from the NIH, Boston and Telethon Biobank blood samples using the Qiagen Puregene kit according to the manufacturer’s instructions. Saliva was collected in an Oragene kit and was prepared using prepIT.L2P. DNA was extracted from immortalized cell lines using the 5 Prime ArchivePure kit.

### DNA Sample Handling and Library Preparation

Quality control of input genomic DNA samples were conducted by visual inspection for discoloration and/or presence of precipitants. Genomic DNA quantitation was performed using a fluorescence dye-based assay (PicoGreen dsDNA reagent) and measured by a microplate reader (Molecular Devices SpectraMax Gemini XS) before normalization to 20 ng/uL. Normalized gDNA samples were added into wells of a Covaris 96 microTUBE plate at 55 uL volume and sheared using the Covaris LE220 Focused-ultrasonicator with settings for targeting a peak size of 410bp (PIP: 450 W, Duty Factor: 18%, Cycles per burst: 200, Time: 60s). Sequencing libraries were generated from 1,000 ng of fragmented DNA using the Illumina TruSeq DNA PCR-Free HT Library Preparation Kit with minor modifications for automation on a Hamilton STAR Liquid Handling System. Adapters for ligation used either TruSeq DNA CD Indexes or IDT for Illumina TruSeq DNA UD Indexes (96 Indexes, 96 Samples). Library size distribution and absence of free adapters and/or adapter dimers was assessed by automated capillary gel-electrophoresis (Advanced Analytical Fragment Analyzer). Library yield and concentration (in nM) was determined by qPCR quantitation using the KAPA qPCR Quantification Kit on a Roche Light Cycler 480 Instrument II.

### Library Clustering and Whole Genome Sequencing

After qPCR quantitation of sequencing libraries, normalization of libraries to 2.2 nM was performed into a working 96 well plate by automation on a Hamilton STAR Liquid Handling System. Libraries were clustered as single lane per single flowcell for germline tissue-derived samples on an Illumina cBot2 using the HiSeq X PE Cluster Kit and a HiSeq X Flow Cell v2.5 before sequencing on an Illumina HiSeq X System with 151+7+151 cycle parameters using HISeq X HD SBS Kit reagents.

DNA samples were sequenced with Illumina HiSeq 3000 platform in Dec 2017 and Illumina NovaSeq 6000 in Feb 2020 with paired-end reads of 150 bp with coverage of ∼35x at the Sequencing Center of the Uniformed Serviced University of the Health Sciences in Bethesda, Maryland. Fastq files were aligned to reference genome hg38 via BWA 0.7.17^33^. Duplicates were masked and sorted with Picard 2.22.2. The bases of reads were recalibrated, and reads were re-aligned against known INDELs and SNPs in 1000-genomes, and variants and small INDELs were jointly called with GATK 3.8.1 by following the best practices of GATK variant calling^34^. These data processes were carried out on the NIH HPC Biowulf cluster. The SNVs detected by GATK variant calling were imputed for missing genotypes in areas of poor sequence coverage and phased with the TOPMED imputation server at https://imputation.biodatacatalyst.nhlbi.nih.gov/. The SNVs were annotated using ANNOVAR version 2019-10-24^35^. Subsequent analysis was performed on non-synonymous SNVs (frameshift deletion/insertion, non-frameshift deletion/insertion, startloss, stopgain, stoploss, and non-synonymous).

### Quality assessment of the demographic and genotyping data

Relatedness of the samples was checked using the King package^36^. Four hundred seventy-three unrelated individuals with WBS were used for the subsequent analysis. The sex reported in research records for the 473 phenotyped participants was validated with reads mapped to chromosomes X and Y. Overall genomic variation within the cohort, generated with the 142,829 SNV genotyping matrix, was assessed with a principal component analysis and visualized in a plot of principal components 1 and 2 (PC1-2) using the “bigstatsR” package^37^. By using clustering information of the individuals, as shown in Figure S1, along with available self-reported race/ethnicity data as a proxy for continental-level ancestry we imputed missing race/ethnicity data. The race/ethnicity-linked clusters in Figure S1 are similar to those generated by the larger 388,377 participant UK Biobank study^13^, suggesting appropriate representation of genotypes.

### Extreme phenotyping of individuals with WBS

Participants with WBS were classified based on severity of their SVAS into four groups: 1) clinically significant/surgical as defined by a history of surgical intervention in the supravalvar aorta (“surgical SVAS”), 2) mild-to-moderate (defined as presence of any SVAS for which surgery was neither recommended nor performed), 3) no SVAS, and 4) unclassified. These groupings were used because although medical records were used whenever possible to confirm parental report, they were not available in all cases and parental recall of surgical intervention was generally considered more reliable than more quantitative metrics such as gradient. Because the degree of SVAS may increase over the first few years of life, we required that the participant be at least 3 years of age to be listed as “no SVAS”. Consequently, an additional category of unclassified participants who were either too young to classify as “no SVAS” or did not have adequate data for the clinician to confidently assign the phenotypic designation was created. Our modifier evaluation focused on comparisons of those with extreme phenotypes, that is, those with surgical SVAS (n=88) and those with no SVAS (n=137). We then assessed for any differences in variant burden, which is defined as sum of 0s, 1s and 2s for genotypes 0/0, 0/1 and 1/1, respectively, for a set of variants of an individual, among the 100,697 autosomal non-synonymous SNVs in the 225 individuals with the extreme phenotypes based on research cohort membership (Boston, NIH, Nevada-Toronto, Telethon), chromosomal sex (XX, XY), sequence batch (year 2017, year 2020), or sample type (blood, saliva, immortalized cells) with separate Wilcoxon tests.

### Variant screening

To search for genetic modifiers in SVAS outside the 7q11.23 region, we expanded the steps used in our previous whole exome study^11^ in two parallel steps for screening both common non-synonymous variants (AF > 5%) and less-frequent/rarer non-synonymous variants (less frequent variants) (AF = 0.5% - 5%) from surgical SVAS and no SVAS participants. To identify common variants most likely to influence outcomes, we focused on variants with 5% or greater AF difference between the surgical SVAS and no SVAS groups with CADD phred score (CADD score) > 10. The selected variants are expected to be the most influential for inferring biological processes that differ between the extreme phenotypes. This approach is similar to the strategy widely used in single-cell RNAseq studies^38^. For the less frequent/rarer variants, we performed two analyses: the first focused on variants with CADD > 10 and present in at least 1% of the individuals with surgical SVAS and none of the individuals without SVAS; we also performed the opposite assessment (CADD > 10, variants in >1% of those with no SVAS and none of those with surgical disease).

### Pathway enrichment using mySigDB

Genes with the selected SNVs were loaded to the v7.4 mySigDB database at http://www.gsea-msigdb.org/gsea/msigdb/annotate.jsp for enrichment in 50 H:hallmark gene sets and 2922 C2 canonical pathways, including CP:BIOCARTA, CP:KEGG, CP:PID, CP:REACTOME, CP:WIKIPATHWAYS. Enriched pathways were selected based on the hypergeometric p-value after correction for multiple testing with FDR q-value threshold set at < 0.05. Pathways with eight or fewer overlapping genes were excluded from further analysis.

### Association of SVAS severity with enriched pathways

We used the RQT R package^32^ and SKAT/SKAT-O in the RVtest package^29^ to test the association of SVAS severity with each of the enriched pathways. The response variable for surgical SVAS and no SVAS was coded as 1 and 0, respectively. Covariates included the genotype matrix for variants in each pathway and chromosomal sex. Pathways are considered significant with FDR-value < 0.05 by RQT test, and SKAT or SKAT-O test. Pathways with similar biological functions were aggregated and represented by the pathway with the highest number of genes. Most of the genes are present in only one of the aggregated pathways.

### Exploring the GWAS catalog with genes of interest

We downloaded the GWAS catalog from the NHGRI-EBI file: gwas_catalog_v1.0.2- associations_e104_r2021-09-23.tsv (https://www.ebi.ac.uk/gwas/) which contains 4,462 GWAS studies on 5,642 diseases/traits. After excluding studies without reported gene(s), we generated a table containing 27,281 genes reported in 4,583 diseases/traits. We then pruned the GWAS list to include only studies that had assessed phenotypes related to the aorta (n=13) and looked for overlap between our list of genes on modifier pathways and those that had been reported in the published GWAS.

### Statistics

The Wilcoxon test implemented in JMP16 software (SAS Institute Inc., Cary, NC) was used for all comparisons in the supplemental figures. R software (https://www.R-project.org/) implemented through Rstudio (http://www.rstudio.com) was used for generating the principal component plot. The p-values calculated from pathway enrichment and association tests were adjusted with the Benjamini and Hochberg method.

## Results

### Demographic information

The 473 participants with WBS (236 females, 237 males) were classified into four categories: no SVAS (n=137), mild-moderate SVAS (n=189), surgical SVAS (n=88), and unclassified (n=59). Demographic information for the participants is presented in Table 1. The relative proportions of participants in the no SVAS, mild-moderate SVAS and surgical SVAS categories are similar to those previously reported in the literature^9, 11, 12^ The median age at the last phenotyping event was 9 years, with an interquartile range from 4 to 18 years. Based on self-report and PC1-2 based imputation for those missing self-identified race/ethnicity, 421 of 473 individuals are of primarily European ancestry, 14 have African ancestry, 5 are of Asian ancestry, while the remaining 33 individuals represented in orange in the PC1-2 of Figure S1 are likely an admixture of European, Asian, and Latine/Admixed American ancestry. The median ages of the four groups: no, mild-moderate, surgical SVAS and unclassified were 13, 7, 10 and 3 years, respectively. The percentage of surgical SVAS in each of the four cohorts: Boston, Telethon in Italy, NIH and Nevada-Toronto ranged from 19% – 25%.

**Table 1.** Demographic information for the 473 participants with WBS in the study. There is no difference in the ratio of number of females to males in the surgical SVAS, mild-moderate SVAS, and no SVAS group (p- value = 0.1 by χ^2^ test). However, the ratio in the surgical SVAS and no SVAS groups is significant with p-value = 0.047.

| Variable |  | All Patients<br>n=473 | No SVAS<br>n=137 | Mild SVAS<br>n=189 | Surgical SVAS<br>n=88 | Un-classified SVAS<br>n=59 |
| --- | --- | --- | --- | --- | --- | --- |
| Sex | Female | 236 | 73 | 98 | 35 | 30 |
|  | Male | 237 | 64 | 91 | 53 | 29 |
| Race | European | 421 | 119 | 170 | 80 | 52 |
|  | Asian | 5 | 3 | 0 | 2 | 0 |
|  | African | 14 | 7 | 5 | 1 | 1 |
|  | Mixed | 33 | 8 | 14 | 5 | 6 |
| Age (yr) | Median | 9 | 13 | 7 | 10 | 2.6 |
|  | Range | 0.01 - 62.6 | 3.73 - 62.6 | 0.01 - 46 | 0.1 - 45 | 0.12 - 60 |

To gain power for discovery, our analysis focused on the data from the 225 individuals with extreme SVAS phenotypes: either surgical (n=88) or no SVAS (n=137). Consistent with previous reports^39^, SVAS severity was greater in males (p=0.047 by χ^2^ test). Each of the ancestry and sex-based subgroups had ratios of 1.2-2.1 individuals with no SVAS to each person with surgical SVAS. The only exception to this was the African ancestry subgroup in which seven had no SVAS and one had surgical SVAS, leading to a 7:1 ratio of no SVAS: surgical SVAS.

In the extreme phenotype cohort, non-synonymous variant burden did not vary by sample collection (Figure S2A, p=0.36), chromosomal sex (Figure S2B, p=0.34), sequencing batches (Figure S2C, p=0.46), or sample type (Figure S2D, p=0.21). We noticed, however, that eight individual samples in the Nevada- Toronto and NIH collections exhibited increased variant numbers. Those eight were mixed in terms of sample type, year of sequencing and chromosomal sex but all eight belonged to the PC 1-2 cluster ascribed to those of African ancestry. Relatively increased variation is a well-known feature of the genomic structure of this subgroup^40^. Because our analysis relies on differences in AF between members of the two extreme phenotype groups, skew in alleles related to ancestral background (in this case 7 in the no SVAS group and only one in the surgical SVAS group) could be conflated with disease outcome.

As such, we performed our subsequent analysis using the 217 individuals (87 with surgical SVAS and 130 with no SVAS) without skew. Comparisons of these findings (n=217) to the findings when the 8 individuals with African ancestry (1 surgical SVAS, 7 no SVAS) were included (n=225) will be discussed later in the text.

### Variant screening

We next focused our attention on the variants most likely to influence disease outcomes: variants with higher CADD_phred score (CADD > 10) and at least 5% difference in allele frequency (AF) between the surgical (n=87) and no SVAS (n=130) groups (Figure 1). This process eliminated about 99% of the original 88,663 variants. Of note, the few variants with extremely high CADD_phred score (CADD > 30), showed little difference in AF between the extreme phenotype groups and may influence phenotype(s) other than SVAS. The maximum difference in AF between the extreme phenotype groups is less than 20%.

**Figure 1.**
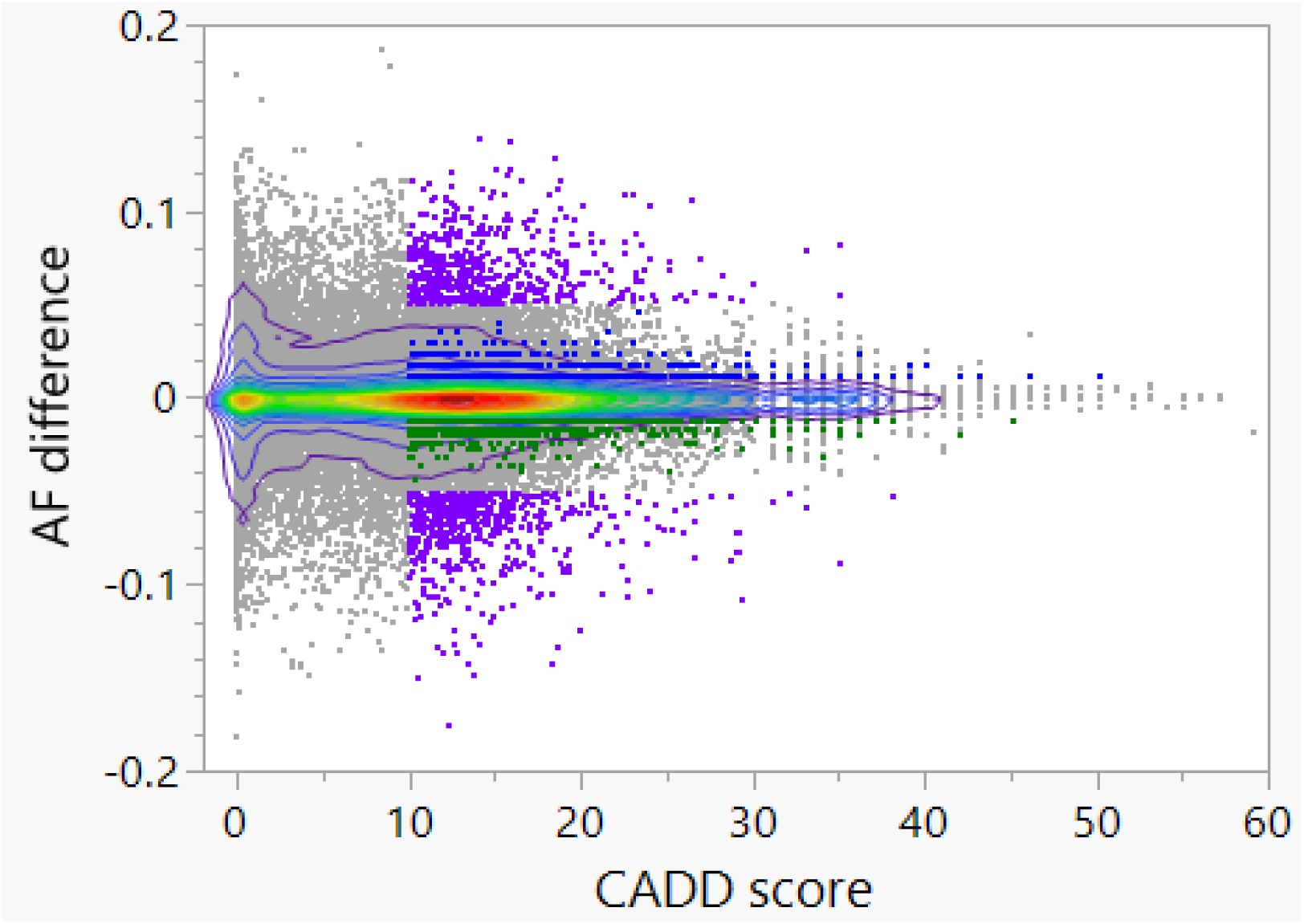
Plot of CADD score vs AF differences between the surgical SVAS (n=87) and no SVAS (n=130) groups for 88,663 autosomal non-synonymous variants. 1064 common variants (914 genes) in purple are selected with > 5% AF difference between the surgical SVAS and no SVAS groups and CADD score > 10. 861 variants (816 genes) in dark green are selected with 0% AF of the surgical SVAS group, > 1% AF of the no SVAS group and CADD > 10; and 1074 variants (995 genes) in blue are selected with > 1% AF of the surgical SVAS group, 0% AF of the no SVAS group and CADD > 10. The area in orange color indicates the highest density of the variants. The outward purple curve is the 90^th^ percentile in the 2-dimensional plot.

After filtration of the primary set, 1064 non-synonymous variants in 914 genes (1.2% of the original variants) remained. The median AF of the 1064 variants in the no SVAS and surgical SVAS groups is 0.28 and 0.29, respectively. Among the 1064, 15 SNVs were stopgain, stoploss, and startloss (see Table S1). Of the 914 genes, 792 have 1 variant each; 104 genes carry 2; and *ZAN*, *CDH23* and *ZNF568* have 5 common variants each. The majority of the variants identified through this approach have a baseline AF > 5% in non-Finnish European (NFE) cohorts in the gnomAD database, meaning they are common variants. No differences in the per-individual burden of the 1064 variants were observed by collection location (Figure S3A, p=0.95), chromosomal sex (Figure S3B, p=0.07), sequencing batch (Figure S3C, p=0.52), sample type (Figure S3D, p=0.89) or SVAS status (Figure S3E, p=0.70) at the 0.05 significance level.

### Pathway enrichment with the 914 genes with 1064 common variants

We hypothesized that variants present at higher AF difference in the surgical SVAS vs the no SVAS groups may be part of the same pathways and that the collective impact of those variants might impart a large influence on physiologic or cellular functions and signaling/communications in cells and tissues. To identify pathways with an increased burden of candidate variants, we performed pathway enrichment using the 914 genes from the 1064 variants; this identified 44 pathways (Table S2). The top pathways include the set of extracellular matrix (ECM) involved genes reported by Naba et al. as the “matrisome”^41^, sensory/olfactory signaling, innate immune system, metabolism of lipids, and ciliopathies.

### Association tests of common variants in pathways with SVAS severity

We next tested each of the 44 enriched pathways for association with SVAS severity. The results from the three methods: RQT, SKAT and SKAT-O are in Table S3. The test results from both SKAT and SKAT-O are similar. Thirty-nine of the 44 pathways met the cutoff of FDR < 0.05 on both the RQT test and the SKAT or SKAT-O test. The 39 pathways shown in Figure 2 were ranked by the FDR-values from the enrichment. Some overlap exists across the 39 pathways, with clear involvement of numerous extracellular matrix pathways, as well as those involving the immune system (adaptive and innate), metabolism of lipids, olfactory (g protein, sensory and perception), and transport of small molecules near the top. Some variant-affected genes, for example in the ECM pathways, are also present in other pathways, including the PI3KAKT pathway, implying interaction among the pathways. In contrast, no gene in the “matrisome” extracellular matrix set is present in ciliopathies, the adaptive immune system or the olfactory transduction pathway. Based on this observation, we then manually consolidated the 39 original pathways to 12 key pathways by grouping pathways with similar functions and overlapping genes, taking as the representative pathway the one with the greatest number of variant-affected genes (shown in Figure 2 with consolidated pathways separated by vertical lines). The 12 representative pathways thus identified include: ECM, sensory/olfactory signaling, innate immune, developmental biology, polymerase II transcription, metabolism of lipids, transport of small molecule, ciliopathies, adaptive immune, PI3KAKT, disease of metabolism and endocytosis.

**Figure 2.**
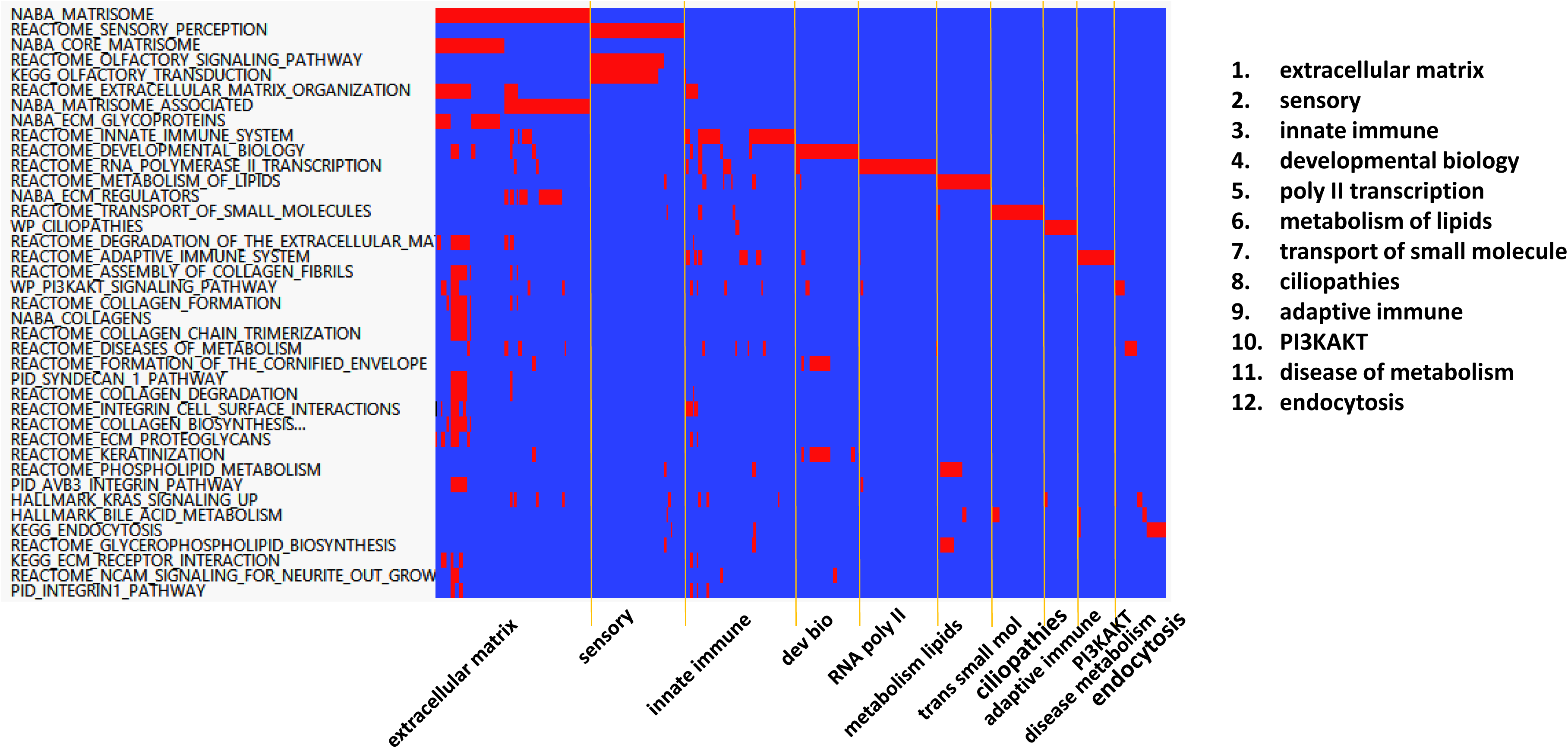
Aggregation of 39 pathways with 360 genes with common variants to 12 pathways. The table was ordered with the most significant enriched pathways on the top.

### Association of SVAS severity with less frequent non-synonymous variants in pathways

As with the common variant analysis, we suspected that less frequent non-synonymous variants might also be concentrated in relevant pathways and may contribute to extreme phenotypic outcomes.

Therefore, we aimed to identify pathways affected by less frequent (AF between 1% and 5%) variants using the extreme phenotyping and variant filtration. To identify less frequent variants that differ between the extreme phenotype groups, we maintained the CADD > 10 screen and then filtered for recurrent variants present only in the surgical or the no SVAS groups. We considered the two sets separately.

In total, 1074 less frequent non-synonymous variants in 995 genes were identified (the blue dots in Figure 1) in the AF > 1% in surgical SVAS and AF = 0% in no SVAS comparison. In addition, 861 variants in 816 genes (the green dots in Figure 1) with AF > 1% in no SVAS and AF = 0% in surgical SVAS were noted. The median AF of the two sets of less frequent variants in surgical SVAS and no SVAS groups are 0.011 and 0.012, respectively.

### Pathway enrichment and association tests using the 995 genes with 1074 less frequent variants in 87 surgical SVAS cases

As for the common variant analysis, we performed pathway enrichment using the 995 genes impacted by the less-frequent variants. Variants from 496 of the 995 gene were statistically enriched (FDR <0.05) into 71 pathways (FDR ranking shown in Table S4). In addition to enrichment in ECM pathways, as seen in the common variant analysis, we also observed enrichment in the gene sets for pathways including apoptotic cleavage of cellular protein/apoptotic execution phase, cell cycle/M phase, ciliopathies, developmental biology, mitotic spindle, RHO-GTPASE, and PI3KAKT.

The association tests by RQT and SKAT/SKAT-O yielded significant results for 58 out of the 71 pathways with FDR-value < 0.05. The test results are shown in Table S5. The 58 pathways were once again manually consolidated to identify 16 major pathways with overlapping gene content, as shown in Figure 3. Interestingly, 28 out of the 87 individuals with surgical SVAS possessed at least one gene with a less frequent variant among the 18 genes in the mitotic spindle pathway, and 47 individuals with surgical SVAS exhibited at least one gene with a less frequent variant among the 36 genes in the cell cycle pathway. Three genes (INCENP, KNTC1 and TUBGCP5) are present in both pathways. Fifty-nine out of the 87 individuals possessed at least one gene with a less frequent variant/gene in one or both pathways.

**Figure 3.**
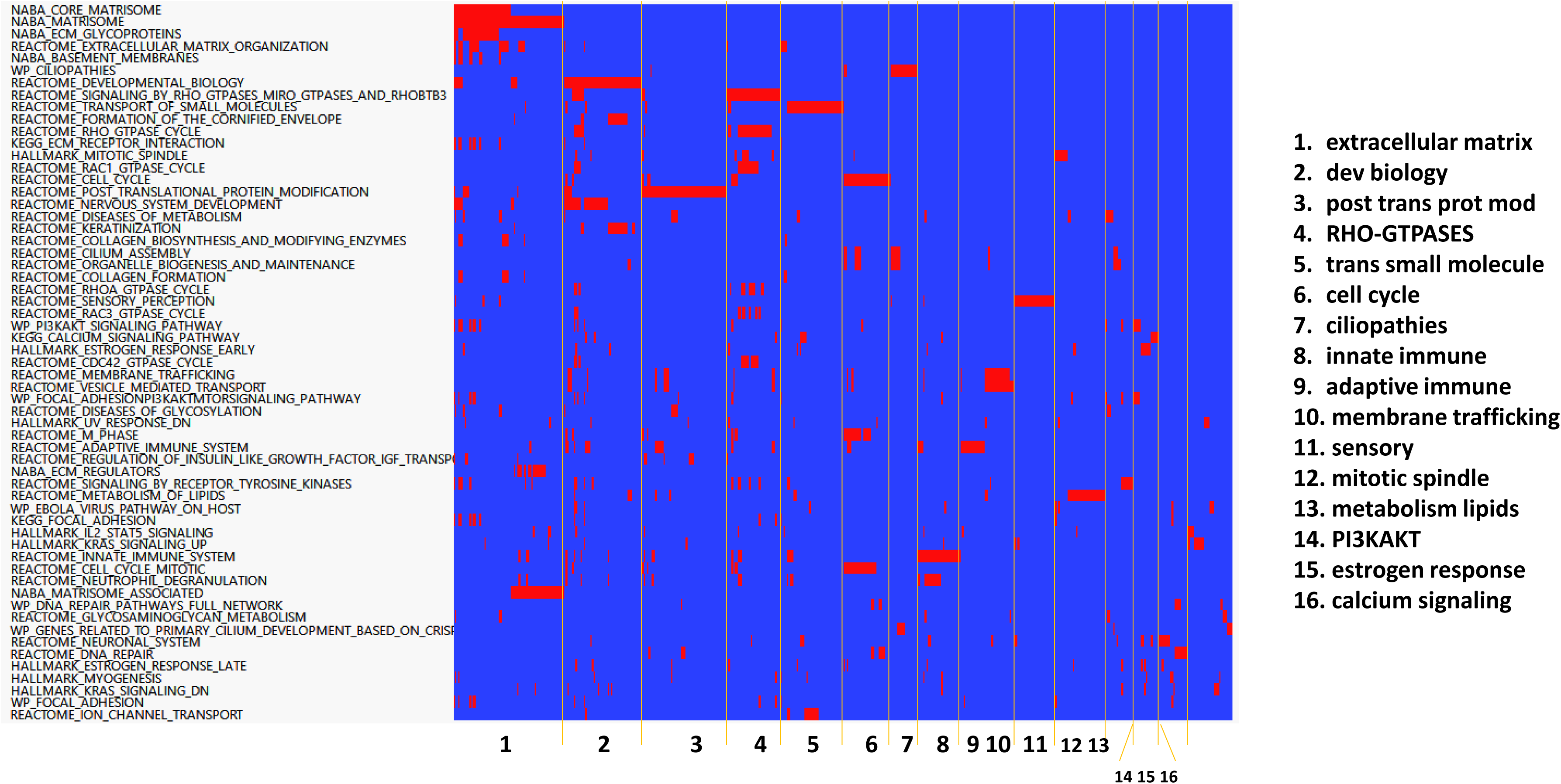
Aggregation of 58 pathways to 16 pathways containing 479 genes with variants selected with AF > 1% in the surgical SVAS group (n=87), AF = 0% in the no SVAS group (n=130) and CADD score > 10.

### Pathway enrichment and association tests using the 816 genes with 861 less frequent variants in 130 individuals without SVAS

Twenty-five pathways showed gene set enrichment with the 816 genes impacted by less frequent variants in the 130 individuals with no SVAS with FDR-value < 0.05. Twenty-three pathways were confirmed by RQT and SKAT/SKAT-O tests at the significance level of 0.05. The test results are shown in Table S6. The 23 pathways were aggregated to 10 major pathways: ECM, post translational protein modification, transport of small molecule, developmental biology, RNA polymerase II transcription, cell cycle, sensory, innate immune, estrogen response and mitotic spindle, shown in Figure 4.

**Figure 4.**
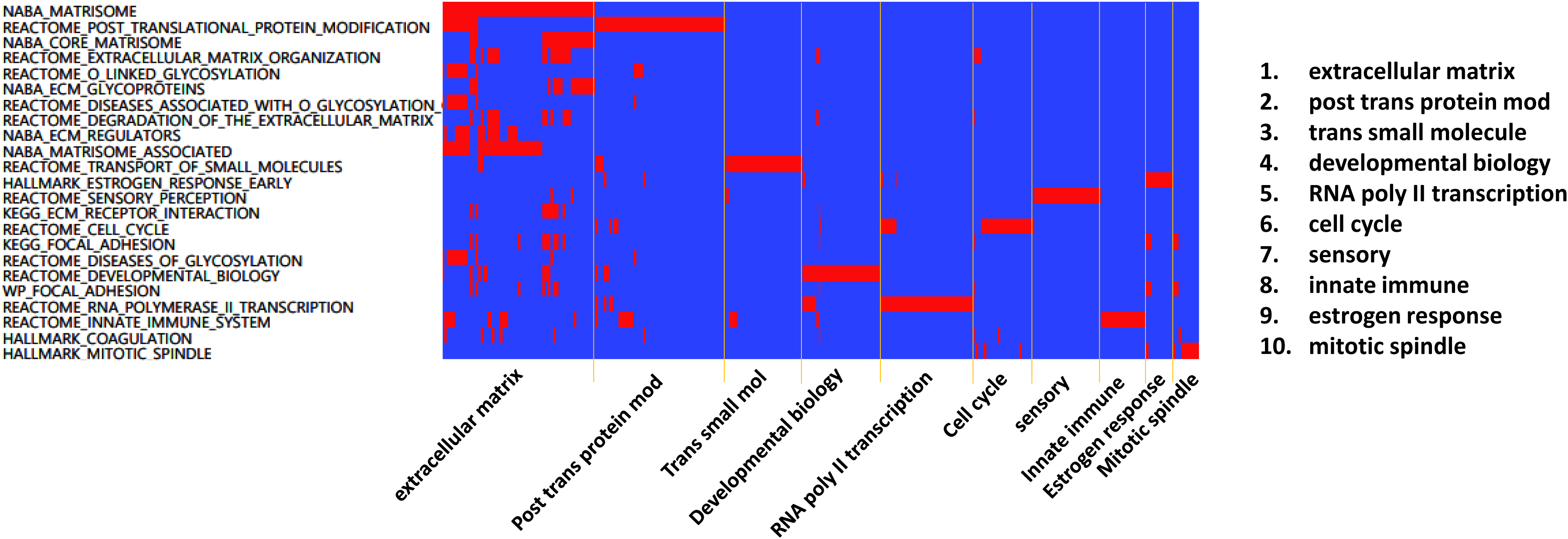
Aggregation of 23 pathways to 10 pathways containing 300 genes selected with AF = 0% in the surgical SVAS group (n=87) and AF > 1% in the no SVAS group (n=130) and CADD score > 10.

### Overlapping the pathways enriched by common variants with two sets of pathways enriched by less frequent variants in the surgical SVAS and no SVAS groups

We then overlapped the three sets of aggregated pathways: 12 pathways derived from genes with common variants, 16 pathways from genes with less frequent variants in the surgical SVAS group, and 10 pathways from genes with less frequent variants in the no SVAS group. The overlaps of the 19 unique pathways are shown in Figure 5. Seven of the 19 pathways are due to the presence of less frequent variants in surgical SVAS or no SVAS: the three pathways in calcium signaling, RHO-GTPASES and membrane trafficking are due to less frequent variants in surgical SVAS, and the four pathways in cell cycle, estrogen response, mitotic spindle and post translational protein modification are due to the presence of less frequent variants in both surgical SVAS and no SVAS. The five pathways in developmental biology, extracellular matrix, innate immune, sensory and transport of small molecule were discovered in all three analyses, indicating that both common and rare variation in these genes contribute to the effect. Four pathways involved in adaptive immune, ciliopathies, metabolism of lipids, and PI3KAKT are present in the set with common variants and the set with less frequent variants in surgical SVAS only. Interestingly, cell cycle, estrogen response, mitotic spindle and protein modification pathways are present only in the two sets of less frequent variants.

**Figure 5.**
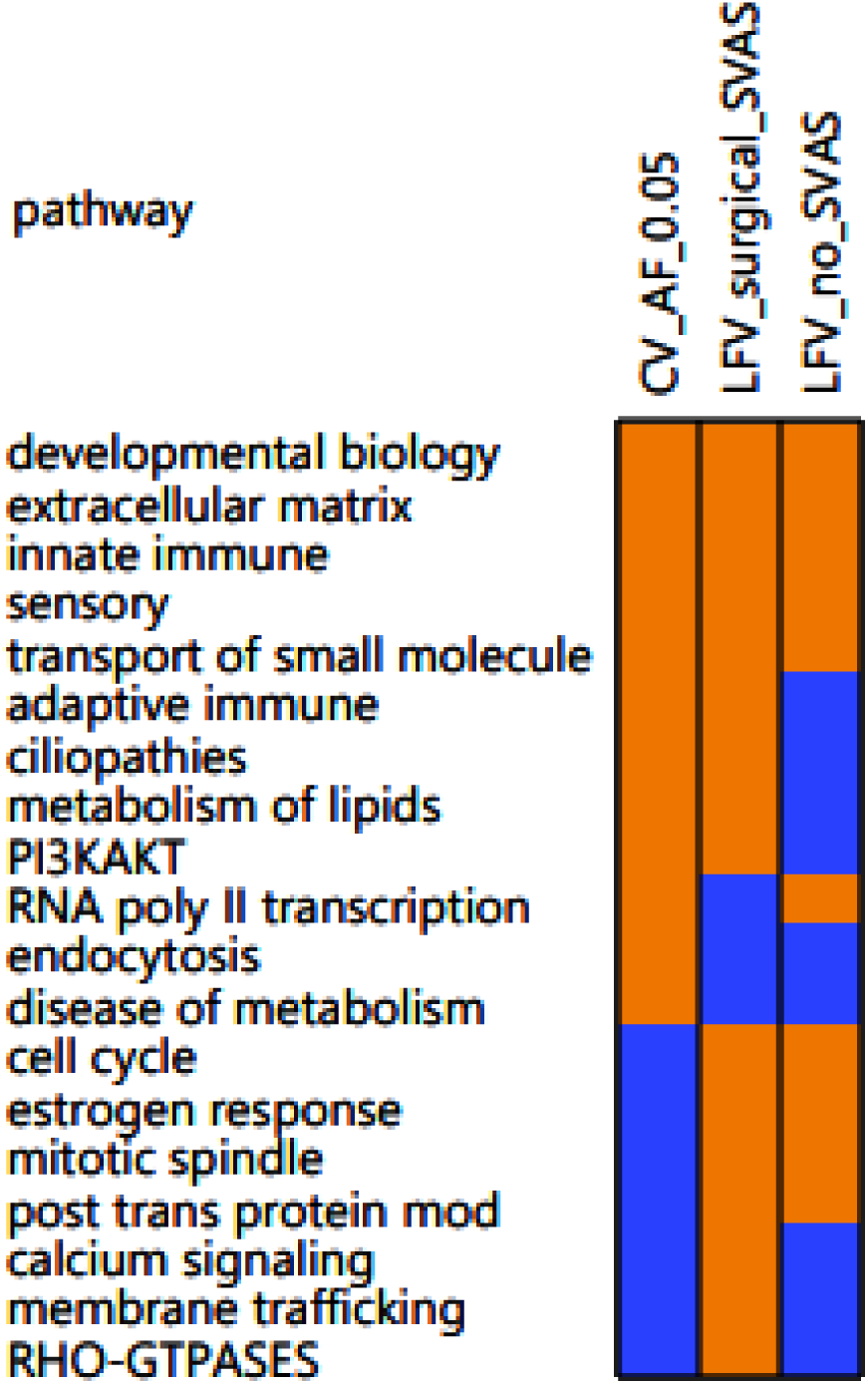
Presence of 19 pathways in three sets of pathways analyses. Presence (in gold) and absence (in blue) of 19 pathways enriched by common variants with AF difference > 5% between the surgical SVAS group (n=87) and the no SVAS group (n=130), “CV_AF_0.05”, by less frequent variants with AF > 1% in the surgical SVAS group, “LFV_surgical_SVAS”, and by less frequent variants with AF > 1% in the no SVAS group, “LFV_no_SVAS”

### Influence of ancestry and phenotypic skew on pathway selection

To determine the impact of skew in a genetic background subgroup, we repeated the three sequential allele frequency-based analyses in the cohort of 225 individuals with WBS, including the eight previously-removed samples with African ancestry. Inclusion of these eight samples yielded a mild impact on the number of significant pathways from association tests for the common variants and little impact on the number of pathways from less frequent variants with AF > 1% in the surgical SVAS and AF = 0% in the no SVAS groups. In contrast, we noted a dramatic increase (see the comparisons in Figure S2) in the number of pathways identified from less frequent variants with AF > 1% in the no SVAS and AF = 0% in the surgical SVAS cases when the eight samples were included. The differences in the pathway sets are driven by variants that are common (AF >5%) in the individuals of African descent whose representation of the extreme phenotypes (surgical SVAS vs no SVAS) is asymmetric, but rare in those of European, Asian, and Latine/Admixed American backgrounds who display similar rates of the two extreme phenotypes. The details of the comparisons are provided in the Supplemental text. Of note, the top pathways remained consistent in both analyses.

### Phenotype-genotype from large GWAS studies in enriched pathways by common and less frequent variants in SVAS

Variants that modify phenotypic outcomes in WBS may perform similar functions in other aortopathies as well. To explore this question, we compared the genes in enriched pathways in this study with genes associated with aortic diseases in the NHGRI-EBI GWAS catalog. In total, 69 genes were reported in 13 GWAS studies on nine aorta-related phenotypes in the catalog, see Table S7. Eleven of these 69 genes, reported in seven GWAS on six aorta traits, are present in 12 pathways in our study. The heatmap of the 11 genes in 12 pathways is shown in Figure 6. Of these, the strongest overlap was seen with genes identified in relation to aortic aneurysm (n=9), while two were ascertained in GWAS of the aortic valve and four were noted in studies of aortic vessel or valve calcification. Four genes were identified in more than one study. Three genes, *IL6R*, *PCSK9*, and *SYMD2*, are of particular interest due to existing clinical studies showing the potential for therapeutic intervention^42–44^.

**Figure 6.**
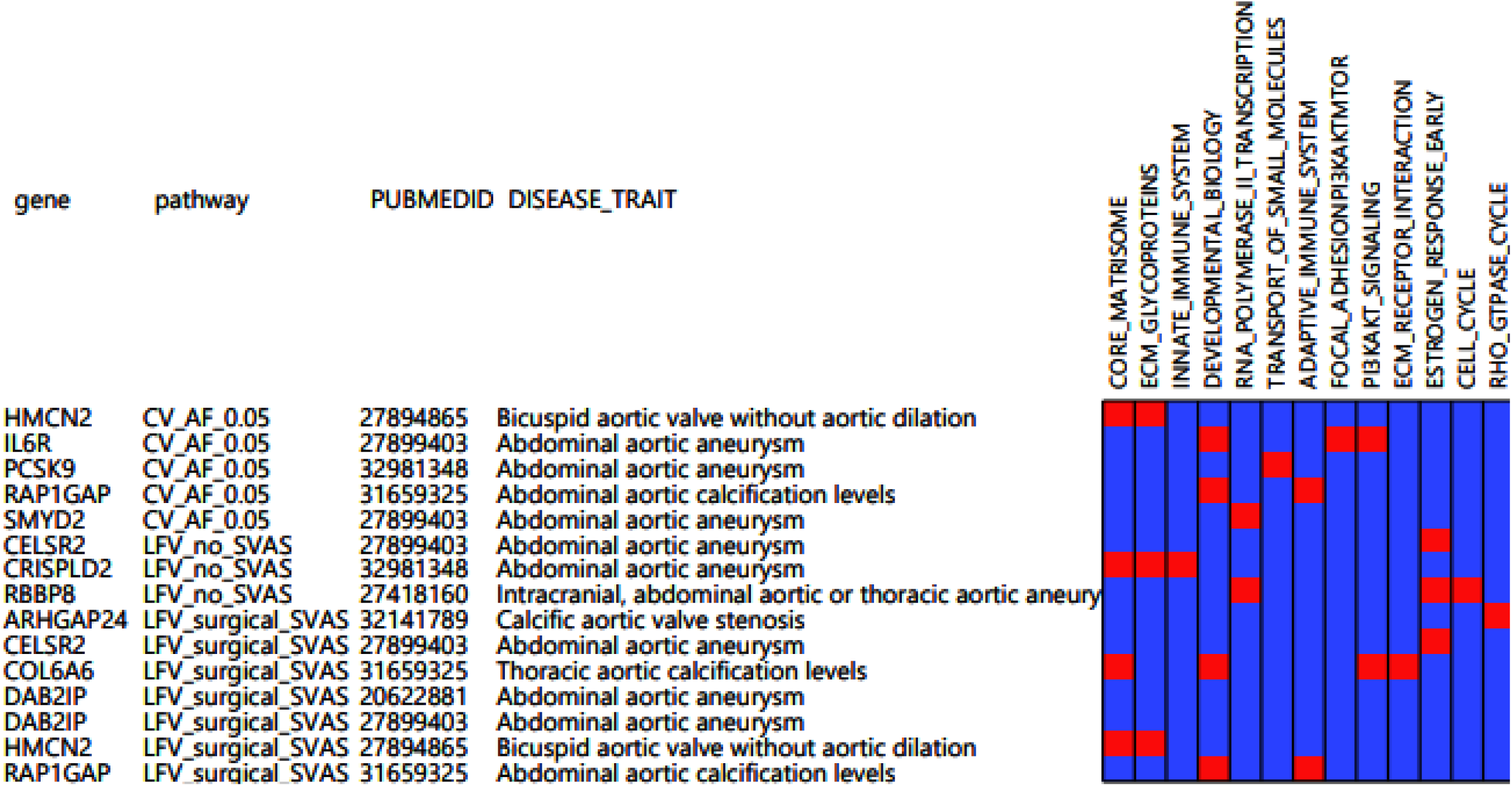
Eleven genes involved in 12 pathways found in our study are also present in seven GWAS studies on six aorta traits.

## Discussion

Although most people with WBS share the same common 7q11.23 deletion, SVAS is not seen in all affected individuals. As such, identification of variation outside of the WBS region that amplifies or mitigates the risk of stenosis may provide both insight into disease mechanism and opportunities for novel therapeutics. However, finding modifiers outside the 7q11.23 region in WBS has been elusive^3, 45^. With the collaboration of four international groups, we were able to carry out the largest genome-wide sequencing investigation in WBS to date. Our analysis centers on the question of whether non-synonymous SNVs (those with CADD score > 10 and large variability in AF for common variants between the two extreme groups) are concentrated in a small number of biological pathways. Our work brings together the efforts of multiple concepts and methods: CADD score in variant effect prediction^27^, variant filtering, similar to selection of the most influential genes for cell type clustering in single cell RNA expression^38^, the pathway approach in transcriptomics analysis^22^, and the association test from population genetics/genomics^23, 24, 29–32, 46, 47^ to provide a framework in which to identify modifier pathways in rare disease cohorts. The analysis starts with the less than 1% of the total number of non-synonymous common variants most likely to impact outcomes (those variants with a marked allele frequency difference between the two extreme groups and potential pathogenicity at the protein level (CADD score > 10). By comparing the list of genes with common and less frequent variants in pathways from our SVAS study to the list of 69 genes found from 13 previous GWAS studies on eight aortic diseases/traits in the NHGRI-EBI GWAS Catalog, we identified several genes that may serve as therapeutic targets based on existing therapies.

In this work, we replicated the findings of Parrish et al^11^ in a larger genome data set. We found association of SVAS severity with pathways in ECM, adaptive immunity, lipid metabolism and GPCR signaling (now referred to under the umbrella of sensory in mySigDB v7.4). The finding that ECM pathway variants may serve as modifiers of elastin insufficiency phenotypes was anticipated. *ELN*, the gene within the WBS locus that is the principal driver of vascular disease in the condition, is an extracellular matrix molecule responsible for aortic mechanics. Deletion of the gene causes arterial stenosis and its duplication in 7q11.23 duplication syndrome causes aneurysm^48, 49^. There are numerous studies showing interactions between elastin and other ECM molecules, in both the development and maintenance of elastic fibers in the aorta^50^. As such, it is not surprising that cumulative small changes in the quantity or quality of other extracellular matrix proteins (specifically pathways in collagens, proteoglycans, integrins and elastic fibers) can synergize to amplify or compensate for elastin-mediated structural and mechanical deficits.

With the larger sample size, we were able to identify associations of SVAS severity with additional pathways including those affecting PI3KAK and KRAS signaling, the innate immune system, and cilia in our common variant analysis. Although these categories may appear somewhat broad, they can inform novel areas of investigation. Mouse experiments have already been performed showing how inhibition of β3 integrins^51^ (a sub-pathway under the ECM category) and the mTOR pathway^52^ (PI3KAK) alter tissue biomechanics and cell function in mouse and iPS models of elastin insufficiency. The data implicating the immune system/inflammation are still developing. Certainly, tropoelastin monomers and elastin degradation fragments are known to influence the immune system^53, 54^. Additionally, our previous study showed an increase in aortic diameter and decreased blood pressure for *Eln^+/-^; Rag1^-/-^* mice that lack B and T cells^11^ and more recently Lin *et al*^55^ showed an influx of monocytes to the area developing stenosis in a new model of elastin insufficiency, the TaglnCre; Eln*^Fl/Fl^*. Further mechanistic details are needed to understand this phenomenon and potentially harness its therapeutic potential. Likewise, additional effort is needed to explain how changes in ciliary function may influence elastin insufficiency outcomes. While cilia are known to play prominent roles in the kidney, retina, and brain^56^, they have only recently been implicated in congenital heart disease^57, 58^, with recent mouse studies identifying abnormal cell migration toward the outflow tract and decreased outflow tract size^59^.

Twelve of the 19 pathways identified in our study were detected by both the common and rarer variant analyses (Figure 5). Seven of them, however, were identified only in the less frequent variant subset.

We strove to include rarer variants because they are thought to underlie much of the missing heritability in many early GWAS^25, 26^, a concern that has been partially addressed in more recent studies by markedly increasing sample size^17^. Unfortunately, the ability to increase cohort size to tens, if not hundreds of thousands, is impossible for a study entirely within a rare disease population. Our method meets this challenge by consolidating rarer variants for analysis on the pathway level, revealing an association of rarer variants in pathways involved with cell cycle/mitotic spindle apparatus and estrogen responsiveness, among others. The cell cycle pathway is intriguing considering the known increase in smooth muscle cell proliferation seen in SVAS lesions^51^. Likewise, estrogen signaling pathways could underlie the reported increase in stenosis severity in males relative to females^11, 39^, that was replicated here in the largest study to date to analyze this phenomenon. Sex hormone effects have also been shown to impact outcomes in other vascular diseases such as vascular Ehlers Danlos syndrome^60^.

Although the variants are individually rare, we showed that 28 of 87 of individuals with surgical SVAS had at least one less frequent non-synonymous variate among the 18 genes in the Hallmark mitotic spindle pathway and 47 of the 87 individuals exhibited at least one less frequent variant among the 36 cell cycle genes in the Rectome cell cycle pathway. These findings suggest that even rare variants within a pathway could cumulatively occur frequently enough to be considered viable modifiers. With growing information from phased haplotypes from long-read sequencing, the cumulative effect of rare and common variants on a haplotype can be better understood and will likely be a driving factor in future genomic research.

Among the five genes with common variants that overlap with previous aortic GWAS, *IL6R*, *SMYD2* and *PCSK9*, stood out due to their appearance in studies specifically evaluating aortic aneurysm^61–65^. *IL6R* is a component of the *IL6* receptor complex and is important for propagating inflammatory signals. Variants in *IL6R* have been linked to increased risk for coronary^66^ and peripheral artery disease^67^, in addition to aortic aneurysm^68^. *SMYD2* also impacts *IL6* levels as well as those of another potent inflammatory molecule TNFa^69, 70^ and is involved in both methylation of genes implicated in aneurysm and in early cell differentiation^71–73^. *PCSK9* was first reported in association with low-density lipoprotein (LDL) levels in large populations^74^, and rare coding variants in the gene are strongly associated with cholesterol levels^17^. Recent studies using drugs targeting either *IL6R* or *PCSK9* have shown efficacy in reducing risk of cardiovascular events^42–44^, with SNV genotype linked to biochemical properties affecting these outcomes. Although SVAS and abdominal aortic aneurysm are two ends of the spectrum of aortic disease, they may in fact share similar modifiers because the two diseases are impacted by the same biologically relevant mechanisms (such as inflammation or aortic size)^75^. It is possible that similar strategies could be considered in people with WBS who have also been shown to have altered immune and metabolic profiles^76^.

### Limitations of our work

In this study we prioritized identification of biological pathways, rather than individual genes, for the association with extreme SVAS phenotypes due to the small sample size. As such, our pathway-level findings are exploratory in nature, and not gene-specific. Incorporating the findings from previous large GWAS studies with the findings from the pathway-level analysis on aortic diseases can be helpful for understanding possible involvement of similar mechanisms in various conditions of the aorta.

Our method is unique in its ability to identify (and replicate) relevant modifier pathways in relatively small groups. However, its emphasis on differences in AF between two extreme phenotype groups offers the potential for confounding in the form of differences that contribute to genetic background (such as ancestry) that are not similarly present in both phenotype groups. Increased variation has been reported in populations with African ancestry in the 1000 genome project^40^. The eight participants with African ancestry in our cohort (7 with no SVAS and 1 with surgical SVAS) exhibited similarly increased variant numbers. Comparison of our 225 and 217 member analyses shows that integration of the 8 samples from the group with African ancestry in the otherwise European ancestry-dominant cohort led to a dramatic increase in the number of filtered variants in one of the rarer variant analyses (Figure S2C). This outcome was driven by unbalanced differences in genetic background on either side of the no SVAS:surgical SVAS comparison. In brief, the seven individuals of African descent who did not have SVAS introduced new variants to the “no SVAS” pool that were common among individuals of this background and triggered the 1% cutoff for the enrichment analysis. Those same variants are uncommon in those with European, Asian and Latine/admixed ancestry. Therefore, because our cohort contained only one person with surgical SVAS who was of African descent, variants associated with genetic background became conflated with those linked to the SVAS phenotype, and they were artifactually retained, affecting the enrichment calculations.

This finding has several important implications. First, larger epidemiological studies are needed to evaluate whether the 7:1 extreme phenotype ratio seen here is representative in larger populations of people with WBS and African ancestry^77, 78^. Currently, the literature contains little focused information about people with WBS of African descent, and further efforts are needed to increase diversity in rare disease reports^79^. Second, although the impact of skew was less obvious in the two remaining analyses, the paucity of non-European ancestral genetic input means that modifiers specific to these backgrounds have not been adequately explored and would benefit from further investigation. Indeed, broad representation is needed to employ robust statistical models that can incorporate samples of multiple ancestries^80^. Clearly, improved engagement, recruitment, and replication in non-European ancestry populations is critical for understanding modifiers of disease outcomes in WBS and other rare diseases. To accelerate discovery in this area, improved funding and international collaborations are needed to engage participants with rare diseases from all backgrounds.

#### Conclusions

This work presents a set of strategies for identifying modifier pathways that contribute to extreme vascular phenotypes in WBS using WGS data with limited cohort size. Both the common and less frequent variant analyses provide new insight into the mechanisms that drive WBS-mediated stenosis outcomes. While ECM pathways like integrins have been previously targeted as part of treatment strategies for elastin insufficiency in mice, the identification of novel pathways such as those affecting inflammation and cell cycle control offer new opportunities for rational therapeutics in this area.

Moreover, the overlap of 11 genes identified in our modifier study (including *IL6R and PCSK9*) with those ascertained through large aortopathy GWAS suggests an overlap in disease mechanism between the groups and highlights the importance of incorporating results in large GWAS studies into studies on rare diseases. Ultimately, this multi-tiered approach has broad applicability. It can be applied to a range of rare diseases and drug trials with small cohorts in which outcomes vary across the population to increase opportunities for pathway-level GWAS. Careful recruitment to maximize inclusion will enhance the lessons learned from these analyses.

## Data Availability

The summary statistics of the non-synonymous variants in no SVAS and surgical SVAS groups will be available upon reasonable request to the corresponding author, Dr. Beth Kozel via.

## Acknowledgement

This work utilized the computational resources of the NIH HPC Biowulf cluster (http://hpc.nih.gov). The graphical abstract was created with BioRender software. The NIH effort was supported by the NHLBI Division of Intramural Research (BAK). BPR and CAM were supported by grants from the Williams Syndrome Association (WSA) and LRO received funding from the Canadian Institutes for Health Research (MOP77720. CBM received support from the National Institute of Neurological Disorders and Stroke (R01 NS35102) and the WSA (WSA 0104 and WSA 0111). CAM and LRO were also partially supported by subcontracts from the grant of R01 NS35102. The Genomic Disorder Biobank of the Telethon Network of Genetic Biobanks was supported by Telethon Italy grant GTB12001G, GM). We would like to thank the individuals with WBS for contributing their DNA samples to this study and the parents who brought their child to the clinics to be evaluated and provided the medical/surgical reports used for phenotyping and answered follow-up questions. Their participation has made the study possible. We also thank Dr. Robert Hufnagel, Dr. Dustin Baldridge, and members of the Kozel lab for discussion and comments.

## Author contributions

D. Liu and B.A. Kozel conceived and designed the analysis. N. Raja, M.D. Levin, E. Bamino, M.F. Bedeschi, M. Digilio, G.M. Squeo, R. Villa, S. Osgood, J.A. Freemen, R.H. Knutsen, B.R. Pober, A.E. Roberts, C.A. Morris, L.R. Osborne, and B.A. Kozel collected the clinical data (patient enrollment and phenotying). W. Resch, C. Dalgard, G. Merla, C.B. Mervis, A.E. Roberts, C.A. Morris, L.R. Osborne and B.A. Kozel contributed data/analysis tools (including data sharing for those not actively involved in phenotyping). Z. Wong, C.J. Billington, Jr, C. Dalgard, C. Alba, and D.N. Hupalo performed DNA sample preparation, sequence data preparation and transferring. D. Liu performed the whole genome sequencing data processing, including alignment, variants calling and imputation. D. Liu and C.J. Billington, Jr performed the statistical analysis. D. Liu and R.H. Knutsen worked on data presentation. D. Liu, C.J. Billington, Jr and B.A. Kozel wrote the paper. All the authors edited and approved the final manuscript. B.A. Kozel supervised the project.

## Competing interests

All authors have completed the ICMJE uniform disclosure form at www.icmje.org/coi_disclosure.pdf and declare: BPR, CAM, and CBM have received funding from the WSA in the past. As a parent advocacy group, the WSA does have an interest in the submitted work but does not stand to financially profit from the findings. Additional government support noted above. No financial relationships with any other organizations that might have an interest in the submitted work occurred in the previous three years; no other relationships or activities that could appear to have influenced the submitted work are noted.

## Data server websites

gnomAD database: https://gnomad.broadinstitute.org/

NHGRI-EBI GWAS catalog database: https://www.ebi.ac.uk/gwas/

mysigDB data: http://www.gsea-msigdb.org/gsea/msigdb/annotate.jsp

TOPMED imputation server: https://imputation.biodatacatalyst.nhlbi.nih.gov/

## Data Availability

The summary statistics of the non-synonymous variants in no SVAS and surgical SVAS groups will be available upon request to the corresponding author, Dr. Beth Kozel via.

## Supplemental figure legend

**Figure S1.**
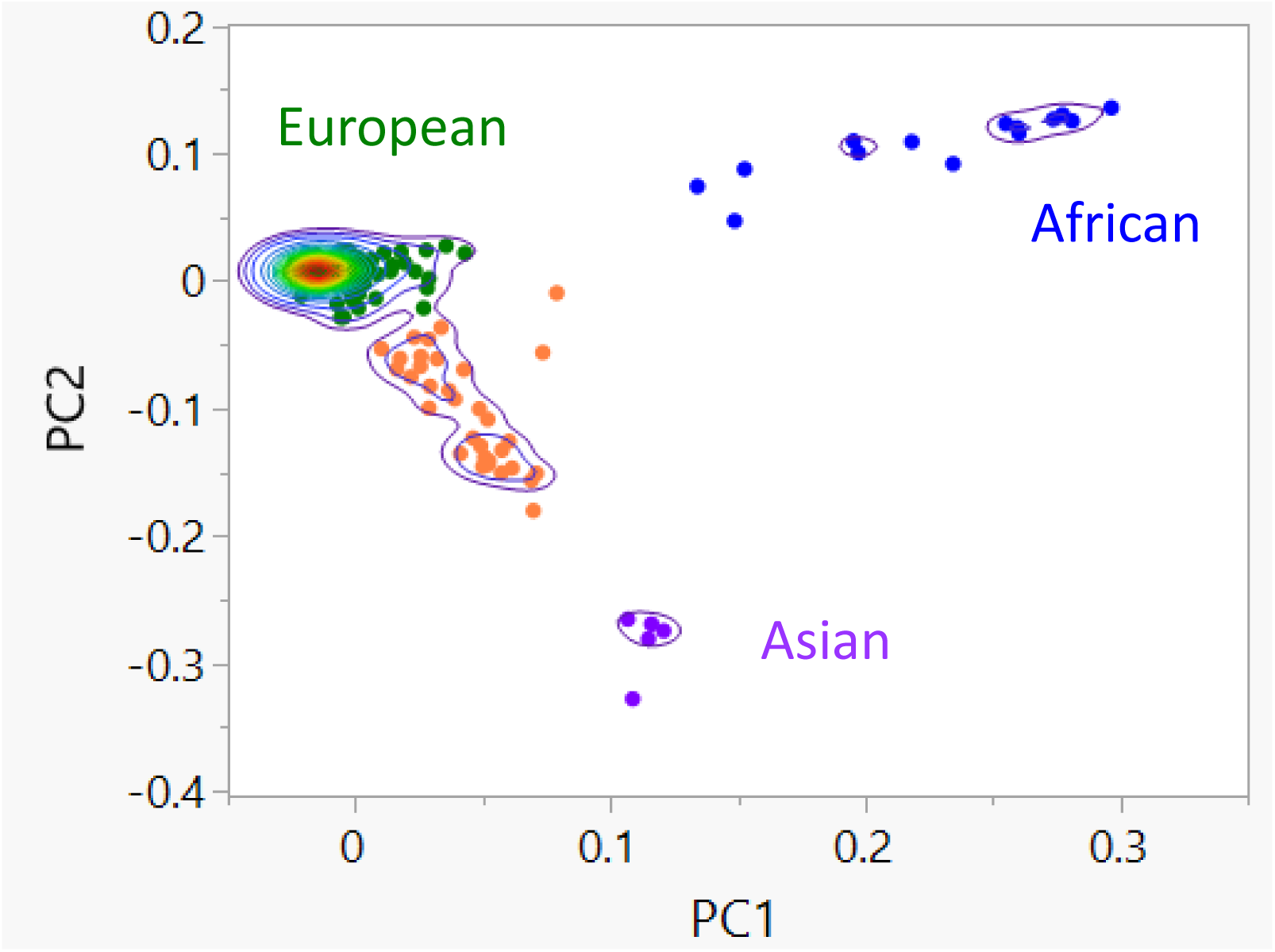
PC1-2 plot generated from the genotypes of 142,829 autosomal non-synonymous variants in 473 individuals with WBS. Missing ethnicity information from medical reports were imputed using clustering information of the individuals with self-reported ethnicity and the individuals having missing self-reported ethnicity. According to our estimate, 421 of the participants with WBS are of European ancestry, 14 African, 5 Asian, and 33 an admixture of European, Asian and Latine. The PC plot was generated using Bigstatsr by Prive et al., Bioinformatics, 2018.

**Figures S2.**
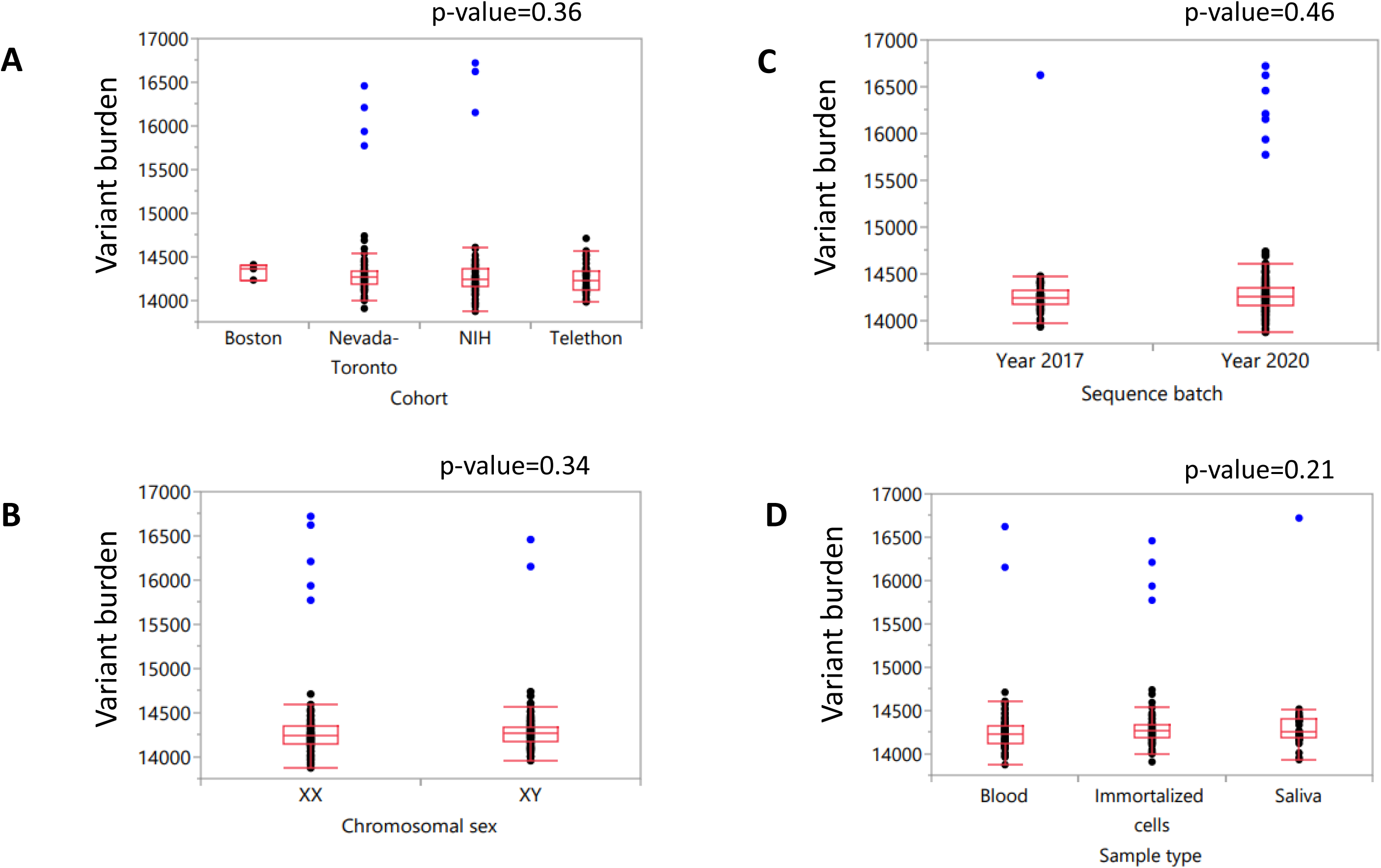
**A-D** Comparison of variant burdens of 100,697 autosomal non-synonymous variants by cohort, chromosomal sex, batch, and sample type, respectively in 225 individuals in surgical SVAS (n=88) and no SVAS (n=137) groups. The eight individuals in blue with African ancestry have increased mutations.

**Figure S3.**
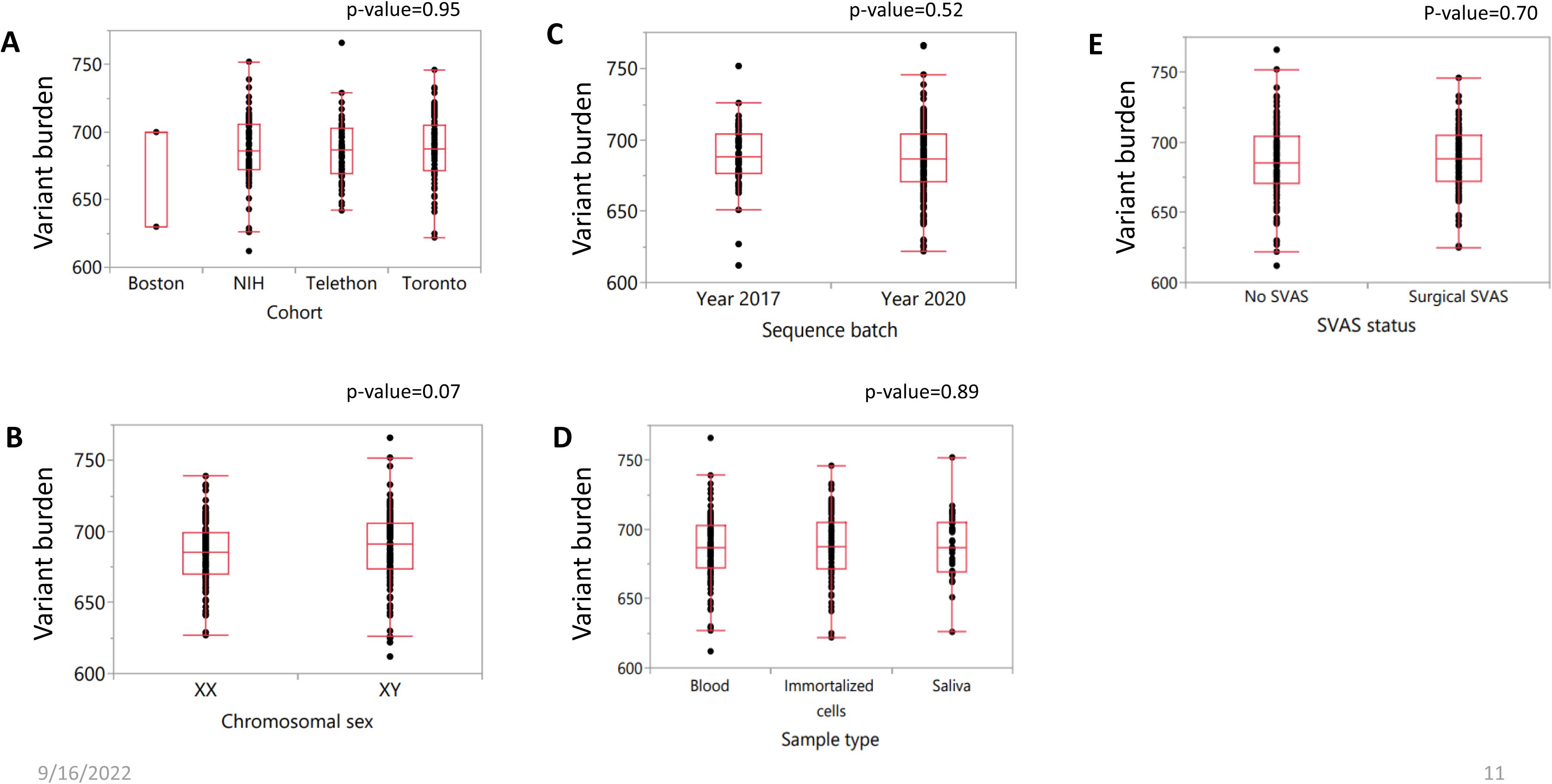
**A-E** Comparison of variant burden of 1064 common variants with AF difference > 5% between the surgical SVAS group (n=87) and the no SVAS group (n=130) by cohort, chromosomal sex, batch, sample type, and SVAS status, respectively.

## Supplemental table legend

Table S1

List of 15 common variants affecting start and stop codons.

Table S2

List of 44 enriched pathways by 914 genes with common variants.

Table S3

List of 39 pathways selected from association tests of RQT and SKAT or SKAT-O and RQT at the level of FDR = 0.05 for common variants with at least 5% AF difference between the surgical SVAS group (n=87) and the no SVAS group (n=130).

Table S4

List of 71 enriched pathways by 496 genes with AF > 1% in the surgical SVAS group (n=87) and AF = 0% in no SVAS group (n=130).

Table S5

List of 58 pathways selected from association tests of RQT and SKAT or SKAT-O at the level of FDR = 0.05 for less frequent variants with AF > 1% in the surgical SVAS group (n=87) and AF = 0% in the no SVAS group (n=130).

Table S6

List of 23 pathways selected from association tests of RQT and SKAT or SKAT-O at the level of FDR = 0.05 for less frequent variants with AF > 1% in the no SVAS group (n=130) and AF = 0% in the surgical SVAS group (n=87).

Table S7

Thirteen GWAS studies on eight aortic traits/diseases in the NHGRI-EBI GWAS catalog. The citations for these studies are provided in the Supplemental text.

## Supplemental text

To determine the influence of the eight individuals with African ancestry (one with surgical SVAS, seven with no SVAS) on the results from the common variants analysis and the less frequent variants analysis, we repeated the analyses previously reported in the manuscript with these eight individuals included (total n=225). We first compared the variant burden of 1049 common variants with AF difference > 5% between the surgical SVAS (n=88) and no SVAS (n=137) groups, including the eight samples with African ancestry by cohort, chromosomal sex, sequencing batch, sample type and SVAS status. The plots are in Figure S1A-E.

**Figure S1.**
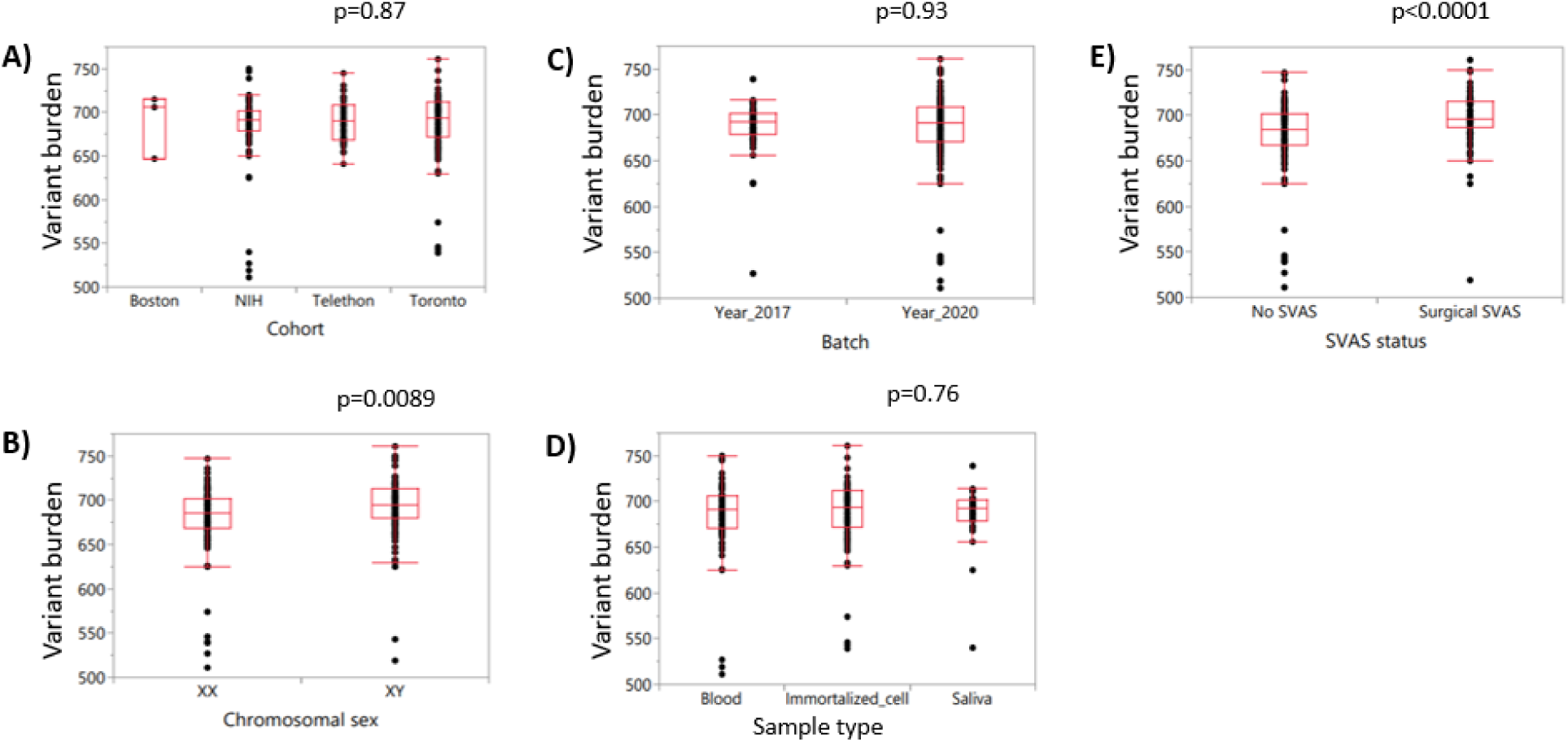
Comparisons of variant burden of 1049 common variants selected with AF difference > 5% between the surgical SVAS (n=88) and no SVAS (n=137) groups and CADD score > 10 by cohort A); chromosomal sex B); batch C); sample type D), and SVAS status F). Wilcoxon tests were used for the comparisons.

We did not find a significant difference in mutation burden by cohort, sequencing batch or sample type, but we did find significant differences by chromosomal sex (p=0.0089, males with higher burden) and SVAS status (p<0.0001, surgical SVAS with higher burden).

We then ran three variant filtering analyses on the group of 225: (A) Common variant analysis AF difference > 5% between the surgical SVAS (n=88) and no SVAS (n=137) groups, and CADD score > 10; (B) Less frequent variant analysis AF > 1% in surgical SVAS and AF = 0 in no SVAS, and CADD score > 10; (C) Less frequent variant analysis AF > 1% in no SVAS and AF = 0 in surgical SVAS, and CADD score > 10. We then compared the three lists of variants from the group of 225 against the three lists from the group of 217 presented in the main text of the paper. For the common variant analysis (A) there are 1049 common variants selected when the eight samples with African ancestry are included and 1064 common variants selected in the n=217 analysis. As shown in the Venn diagram in Figure S2A, the two lists have 878 variants in common, 171 variants are present only when the samples with African ancestry are included, and 186 variants are present only when the samples with African ancestry are removed. For analysis category (B), 1016 less frequent variants are present on both lists; 42 variants are present only in the full sample (n=225), and 58 less frequent variants are only present when the eight samples with African ancestry are removed, Figure S2B. Using analysis category (C), 837 less frequent variants are present in both lists; 516 variants are only present in the full sample (n=225), and 24 are present only when the eight samples with African ancestry are removed, Figure S2C.

**Figure S2.**
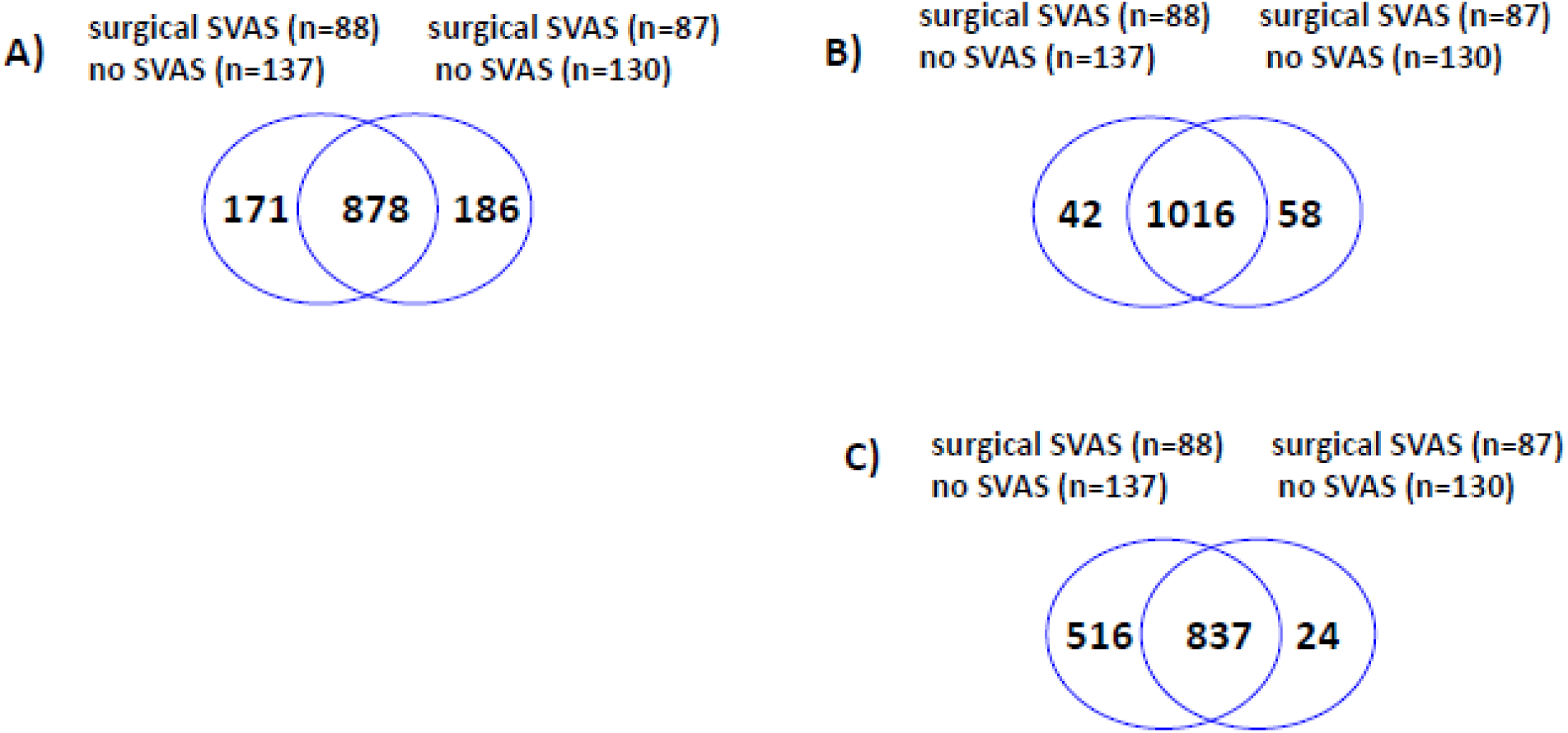
Number of overlapping variants selected from the full sample (n=225, including eight individuals with African ancestry) and the sample removing the eight individuals with African ancestry (n=217), as a function of three sets of criteria: A) AF difference > 5% and CADD score > 10; B) AF > 1% in surgical SVAS, AF = 0 in no SVAS and CADD score > 10; C) AF = 0 in surgical SVAS, AF > 1% in no SVAS and CADD score > 10.

We then checked the variant burden of 1049 common variants in the full sample (n=225), 1058 less frequent variants present in surgical SVAS (n=88) and 1353 less frequent variants present in no SVAS (n=137) across the four groups with ancestry of African (n=8), Asian (n=5), European (n=199), and admixture of Asian, European, or Latine (n=13). The median of variant burden in the 1049 common variants in the African, Asian, European and admixture groups is 540, 630, 695 and 674, respectively. The eight individuals with African ancestry have the lowest variant burden in the common variants, shown in Figure S3A. Within the surgical SVAS group, the median variant burden of the individual with African ancestry is the highest, followed by the two Asian, five admixture, and 80 European respectively, as shown in Figure S3B. Similarly, in the no SVAS group, the median variant burden of the seven individuals with African ancestry is 219, which is the highest; and followed by three Asian, 119 European and eight admixture respectively, shown in Figure S3C. The results indicate that the high variant burden of the seven individuals with African ancestry contributes to over five hundred additional variants after including them in the analysis of less frequent variants present only in the no SVAS group (n=137).

**Figure S3.**
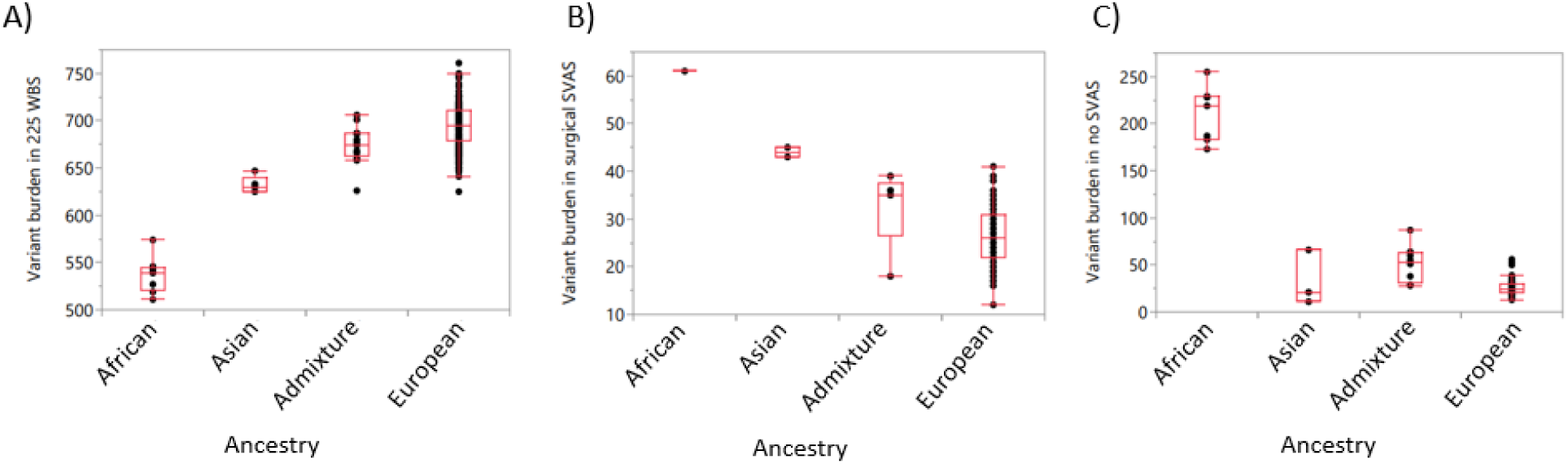
Comparison of variant burden by ancestry of African, Asian, European, and admixture of Asian, European, and Hispanic: A) 1049 common variants in the full sample (n=225); B) 1058 less frequent variants in surgical SVAS (n=88); C) 1353 less frequent variants in no SVAS (n=137).

We also compared the number of overlapping pathways selected from the three analyses between the full sample (n=225) and the analysis without the eight individuals of African descent (n=217): as above A) pathways enriched by common variants selected with AF difference > 5% between the two groups and CADD score > 10; B) AF >1% in surgical, AF = 0% in no SVAS and CADD > 10; C) AF > 1% in no SVAS, AF = 0% in surgical and CADD score > 10. The plots are shown Figure S4. Notably, including the eight individuals with African ancestry does not result in a dramatic difference in number of pathways enriched by common variants, Figure S4A, or less frequent variants in the surgical group, Figure S4B. In contrast, it has the most impact on number of pathways enriched by variants with AF > 1% in no SVAS, Figure S4C: 60 additional pathways are present due to the addition of the eight individuals, as compared to the 25 pathways identified in both cohorts.

**Figure S4.**
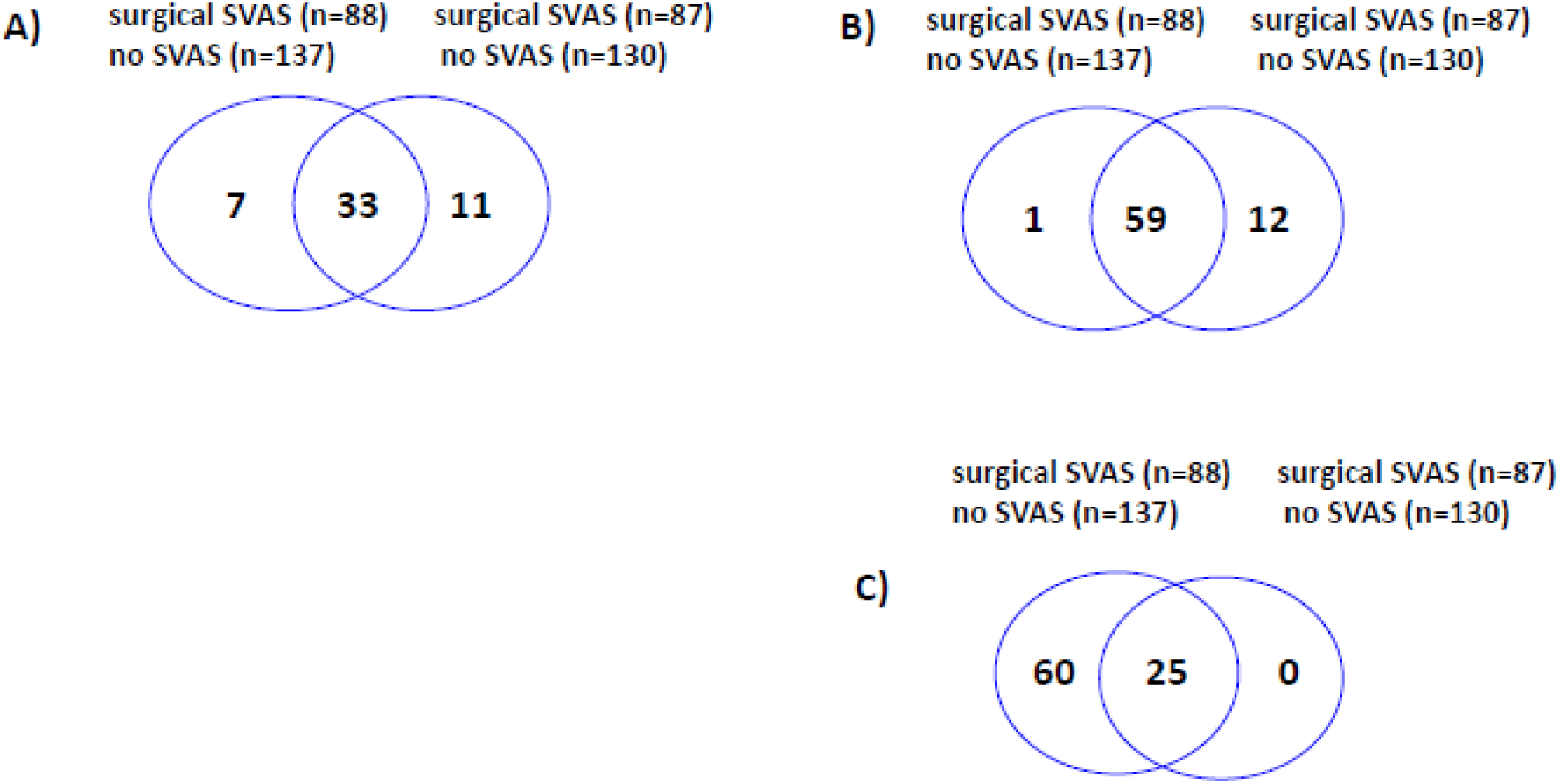
Number of overlapping pathways selected the analysis with the full sample (n=225) and the analysis with the sample excluding the eight individuals with African ancestry (n=217) with three sets of criteria: A) AF difference > 5% and CADD score > 10; B) AF > 1% in surgical SVAS, AF = 0% in no SVAS and CADD score > 10; C) AF = 0% in surgical SVAS, AF > 1% in no SVAS and CADD score > 10.

We then compared the number of significant pathways from the association tests by RQT and SKAT/SKAT-O from two separate analyses: with the full sample (n=225) and the sample (n=217) without the eight individuals with African ancestry. We observed seven additional statistically significant pathways due to inclusion of the eight individuals with African ancestry in the common variant analysis, as well as 25 pathways common to the analyses with and without the eight individuals, shown in Figure S5A. In the less frequent variant analysis of AF > 1% in surgical SVAS, including the eight individuals only yielded one additional statistically significant pathway, and removed 12 pathways identified through the analysis without the eight individuals, shown in Figure S5B. In contrast, In the less frequent variant analysis of AF > 1% in no SVAS, we observed 35 additional pathways from the analysis including the eight individuals with African ancestry. The dramatic difference is due to the fact that the allele frequencies of all 516 variants in Figure S2C are great than 5%, which are common variants.

**Figure S5.**
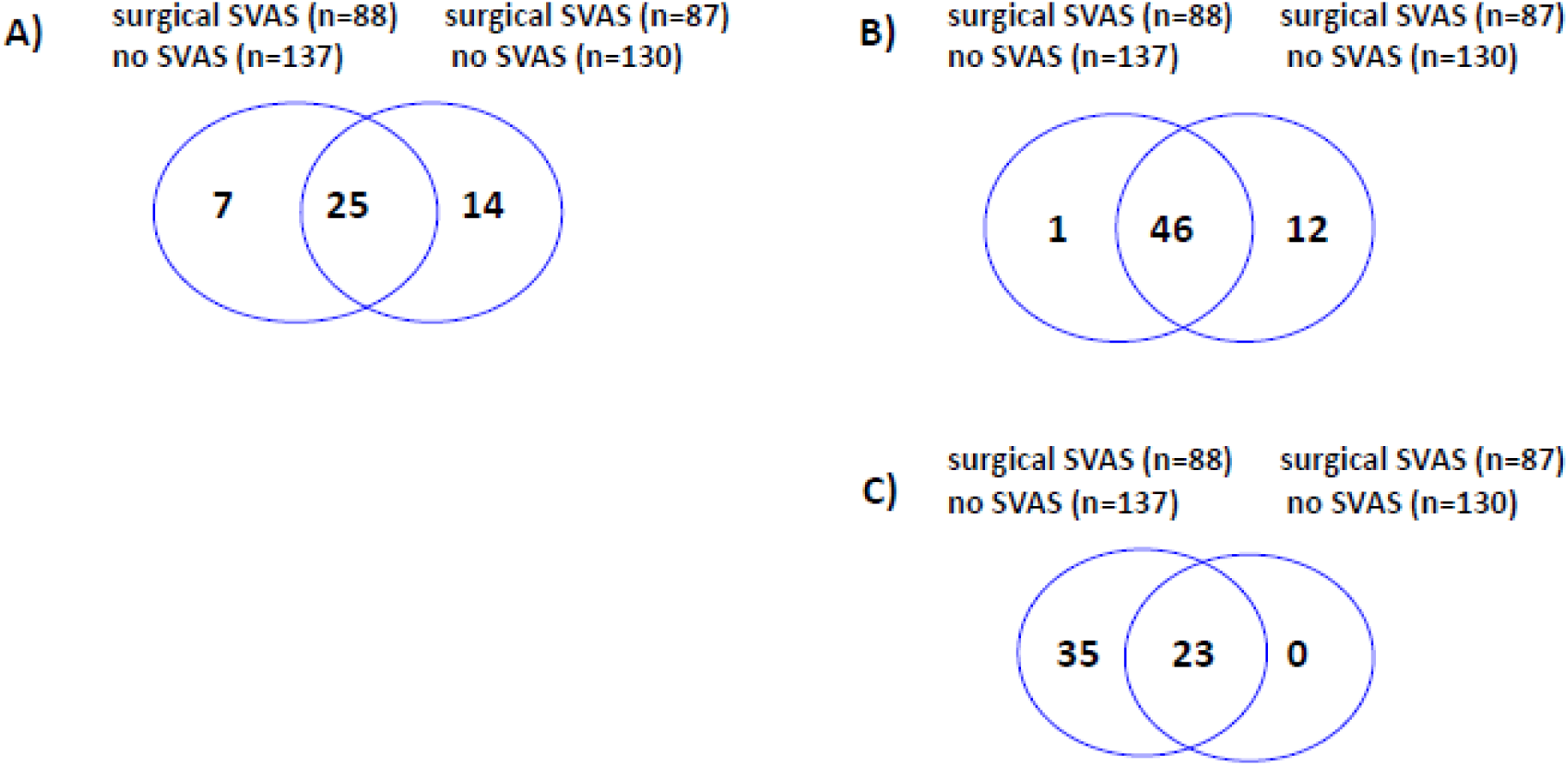
Number of overlapping significant pathways from association tests by RQT and SKAT/SKAT-O with the full sample (n=225) and the sample without the eight individuals with African ancestry (n=217) with three sets of criteria: A) AF difference > 5% and CADD score > 10; B) AF > 1% in surgical SVAS, AF = 0% in no SVAS and CADD score > 10; C) AF = 0% in surgical SVAS, AF > 1% in no SVAS and CADD score > 10.

In summary, we have described the impacts of genetic background of the eight individuals with African ancestry based on three separate comparisons: variant filtering, pathway enrichment, and association tests. The less frequent variant analysis is most altered by the inclusion of the additional samples with African ancestry due to the conflation of ancestry specific common variants with disease outcomes due to the imbalance of SVAS severity in this small cohort (see discussion for additional details). The statistical issues in large GWAS studies with multiple diverse populations have been studied and addressed with mixed models. However, additional methodological developments in study design and analysis for rare disease studies using whole genome sequencing of individuals with diverse ancestral backgrounds are needed.

A summary of the 13 GWAS studies on aortic diseases/traits: ^1–13^ is given in Table S7.

## Notes

### Author Declarations

Institutional Review Board of the National Institutes of Health, the University of Nevada School of Medicine Internal Review Board, the University of Toronto Health Sciences Research Ethics Board, the Boston Children Hospital Internal Review Board, and Fondazione IRCCS Casa Sollievo della Sofferenza Ethics Board gave ethical approval for this work.

